# Overview of effects of motor learning strategies in neurological and geriatric populations: a systematic mapping review

**DOI:** 10.1101/2024.06.19.24309068

**Authors:** Li-Juan Jie, Melanie Kleynen, Guus Rothuizen, Elmar Kal, Andreas Rothgangel, Susy Braun

## Abstract

**Introduction:** Motor learning plays a central role in neurological and geriatric rehabilitation. The wide range of motor learning strategies and increase in evidence can make it difficult to make informed decisions about the use of motor learning strategies in practice. This review’s aim was to provide a broad overview of the current state of research regarding the effects of seven commonly used motor learning strategies to improve functional tasks within older neurological and geriatric populations.

**Method:** A systematic mapping review of randomised controlled trials was conducted regarding the effectiveness of seven motor learning strategies – errorless learning, analogy learning, observational learning, trial-and-error learning, dual-task learning, discovery learning, and movement imagery – within the geriatric and neurological population. PubMed, CINAHL, and Embase databases were searched. The Risk of Bias 2 tool was used to assess bias; additionally, papers underwent screening for sample size justification.

**Results:** Eighty-seven articles were included. Identified articles regarding the effects of the targeted motor learning strategies started around the year 2000 and mainly emerged since 2010. Eight different populations were included, e.g. Parkinson’s, and stroke. Studies were not equally balanced across the motor learning strategies or target groups and overall showed a moderate to high risk of bias. Positive trends regarding effects were observed for dual-tasking, observational learning and movement imagery.

**Conclusions:** The findings show a skewed distribution of studies across motor learning interventions, which have been researched within a variety of populations. Methodological shortcomings make it difficult to draw firm conclusions regarding the effectiveness of motor learning strategies. Future researchers are strongly advised to follow guidelines that aid in maintaining methodological quality. Moreover, alternative designs fitting the complex practice situation should be considered.

---

Motor learning - defined as a relatively permanent change in performance or behaviour^1^ - plays a central role in the rehabilitation of neurological and geriatric rehabilitation.^2–4^ Healthcare professionals, such as physical and occupational therapists, support patients to acquire or relearn a broad range of different motor skills, e.g. walking or reaching, to help them regain independence in activities of daily living (ADLs). ^1–4^ Certain general principles of skill training, like the frequency and specificity of practice and number of repetitions, are now widely recognised as being crucial to effective rehabilitation. In the last two decades, evidence has accumulated to suggest that *how* skills are taught may also be of relevance, ^5,6^ and guidelines now recommend incorporating motor learning strategies (e.g. implicit and explicit motor learning) into treatment approaches to improve rehabilitation success.^7^ However, in many cases, there remains a lack of clear guidance on how different motor learning strategies can best be incorporated (and which can best be used for whom).^7–9^ The vast variety of motor learning strategies from which health care professionals can choose, together with the rapid growth in publications and lack of overview of the effectiveness of these different strategies make it challenging to make informed decisions regarding the appropriate treatment approach and motor learning strategies. Further, many healthcare professionals seem to acquire novel knowledge unsystematically and in a fragmented manner.^9,10^

In order to support clinicians’ decision-making and aid evidence-based implementation of motor learning strategies in their clinical practice, Kleynen and colleagues^11^ developed a practical framework based on the broad distinction between conscious and non-conscious attributes of the motor learning process. This distinction proposes that implicit motor learning targets more non-conscious attributes of the motor learning process, whereas explicit motor learning targets more conscious attributes of the motor learning process.^12,13^ The framework includes seven common motor learning strategies, which have been categorised as promoting more implicit or explicit motor learning: errorless learning, dual-task learning, analogy learning, discovery learning, observational learning, movement imagery, and trial-and-error learning.^11,14^ The framework was informed by practice-based evidence from experts in different fields (e.g. researchers, health care professionals) as well as by research results that underpin these different learning strategies’ working mechanisms. Currently, most evidence regarding the effectiveness of more implicit and explicit forms of motor learning is based on studies using laboratory tasks, e.g. Kal et al.^15^ To support healthcare professionals in making informed decisions about the use of the seven motor learning strategies, more insight into their effectiveness in functional tasks is needed.

Systematic reviews potentially provide therapists with an accessible overview of available evidence to support their decision-making process. However, these reviews are often limited to a single motor learning strategy (e.g. errorless learning) within specific target populations (e.g. pathology- or disease-based), and focus on a single measurement outcome as to allow data pooling or synthesis (e.g., see^16–18^ for excellent examples). In clinical practice, however, therapists treat various populations with a great variety of motor problems (and thus outcomes), rehabilitation needs, and preferences. Therapists therefore may need to switch between strategies, both within and between patients, to provide an optimal learning environment – but lack clear guidance to base this on as more comprehensive overview of the motor learning literature is lacking. This study’s aim was to perform a systematic mapping review of randomised controlled trials (RCTs) to provide a comprehensive overview of the effects across the seven motor learning strategies incorporated in Kleynen et al.’s motor learning framework^11^ for neurological and geriatric populations. In addition, the content of the interventions is described to gain more insight into how therapists could perform the different strategies in clinical practice.

## Methods

This systematic review was conducted in two parts. The first part consisted of a quantitative analysis and focused on mapping the included studies to gain a quick overview of how many were published per (sub)population and per motor learning strategy over time. The second part included a descriptive analysis of motor learning intervention contents and effects, critically appraised in light of the studies’ risk of bias and sample size justification.

### Eligibility criteria

The population included all adults older than 60 and was not restricted to certain disorders. However, to optimise the search strategy, potentially relevant populations were specifically included in the search function (see search strategy). To ensure that the included studies would have direct clinical relevance, these studies’ aim should be a performance improvement in a functional movement task.

We defined a functional task as an activity that individuals perform as part of their daily routine, work, hobbies, or rehabilitation program. The control intervention group (comparator) was not predefined. The eligibility criteria for the selection of studies are presented in Table 1.

**Table 1.** Overview of selection criteria.

|  | **Inclusion** | **Exclusion** |
| --- | --- | --- |
| **Population** | Mean ≥ 60 years | Mean age < 60 years |
| **Intervention** | Errorless learning, analogy learning, observational learning, trial-and-error learning, dual-task learning, discovery learning, and mental imagery | Multimodal training interventions to ensure training effects are due to one of the seven interventions rather than other training modalities or combined interventions |
| | ≥ 1 training session | < 1 training session |
| **Outcome** | Physical movement performance outcome assessed immediately after the intervention (acquisition) and/or at a delayed time point (retention/ transfer) | No motor performance outcome was measured, e.g. only magnetic resonance imaging or electroencephalogram. No serial reaction time outcomes |
| **Study Design** | Randomised controlled trial | Any other non-randomised trial design |
| **Language** | English, German, or Dutch | - |
| **Accessibility** | - | No full text available |

### Search strategy

Two researchers (LJ, GR) searched the databases PubMed, CINAHL, and Embase for randomised controlled trials using the following combination of key search terms: ageing (older adults) OR neurological diseases (stroke OR parkinson OR dementia) AND motor learning strategies (analogy learning OR errorless learning OR trial and error OR discovery learning OR dual-task learning OR action observation OR mental practice) AND Activities of Daily Living (functional tasks). A detailed overview of the search strategy and the search terms used can be found in Appendix A. Additionally, reference tracking of the included studies was performed to identify additional studies.

### Study selection process

#### Identification and screening of studies

Two researchers (GR, LJ) independently screened all retrieved articles from the databases based on the title, abstract, and keywords. After screening, the same two researchers obtained and assessed the full text of eligible articles independently according to the predefined selection criteria. In case of persistent disagreement, a third reviewer (MK) was consulted to reach a consensus.

#### Risk of bias assessment and sample size justification

The Cochrane Risk of Bias 2 (RoB2; ^19^) was used to evaluate five different domains of bias, namely, randomisation, deviations from the intended intervention, missing outcome data, measurements of the outcome, and reporting of results. Based on specific criteria for each domain, an overall risk of bias was determined for every study, ranging from low risk of bias (green) to some concerns (yellow) to high risk of bias (red). Given the large number of studies included, a total of seven assessors (LJ, MK, GR, AR, EK, SB, RS) were involved in rating the risk of bias. To increase the reliability of the ratings, four calibration sessions were organised in which each item was discussed and further specified for the context of this study. Each article was assessed by two independent assessors. In case of disagreements, a third researcher was consulted. Authors of the included studies were not contacted to retrieve missing information. In addition to the standard RoB2 items, one extra item regarding the studies’ sample sizes was added, since appropriate sample size justifications are often lacking or not transparently described.^20^ Studies were also evaluated based on whether an a priori power analysis or other form of sample size justification was described. If sample size justification was described and achieved, this was categorised as ‘green’. If no appropriate size justification was provided or the required sample size was not achieved, this was categorised as ‘red’.

#### Data extraction

The following characteristics were extracted: year of publication, author, number of participants in total and per intervention/control group, population studied, gender, mean age, movement task trained, type of motor learning strategy/intervention, frequency and total duration of supervised practice, movement performance measures, assessment time points, and between-group effects.

#### Data analyses and synthesis

The analysis was divided into 1) a quantitative analysis in which the current available studies were mapped (Q1-3) and 2) a descriptive analysis of the studies’ characteristics, the content of the intervention, and synthesis of the potential effects (Q4). For the quantitative analysis, all eligible articles were included. In the descriptive analysis, to increase the reliability of this study’s conclusion regarding intervention effects, a second selection took place in which studies that scored ‘high’ on RoB2 and lacked (or failed to meet) an appropriate sample size justification were excluded.

As part of the quantitative analysis, a flowchart was presented to visualise the search and selection procedure. Further, the number of included studies per learning strategy over time and the type of patient population per learning strategy were mapped. An overview table per learning strategy was created presenting the risk of bias (low, some concerns, or high), sample size justification (yes/no), population, number of participants, and task trained.

As part of the descriptive analysis, more in-depth information was provided regarding the population (type, group sizes, gender, age), intervention (motor learning strategy(ies), control intervention(s)), duration and frequency, task trained, movement performance measurement, and measurement moments (e.g. immediately after the intervention and, if applicable, also at follow-up). In the last step, we descriptively synthesised between group differences, both in terms of significance and direction of effects.

## Results

### Study selection

The study flow is visualised in Figure 1. In total, 2099 articles were identified. After deleting duplicates and screening the titles and abstracts, 236 articles remained, of which five articles could not be retrieved. The full text of 231 articles was obtained, and after screening, 90 were considered eligible and included for further analysis. Within this sample, there were three occasions in which the data of one single RCT was analysed in two different papers.^21–25^ These papers were counted once, leading to a grand total of 87 studies included in the current review.

**Figure 1.**
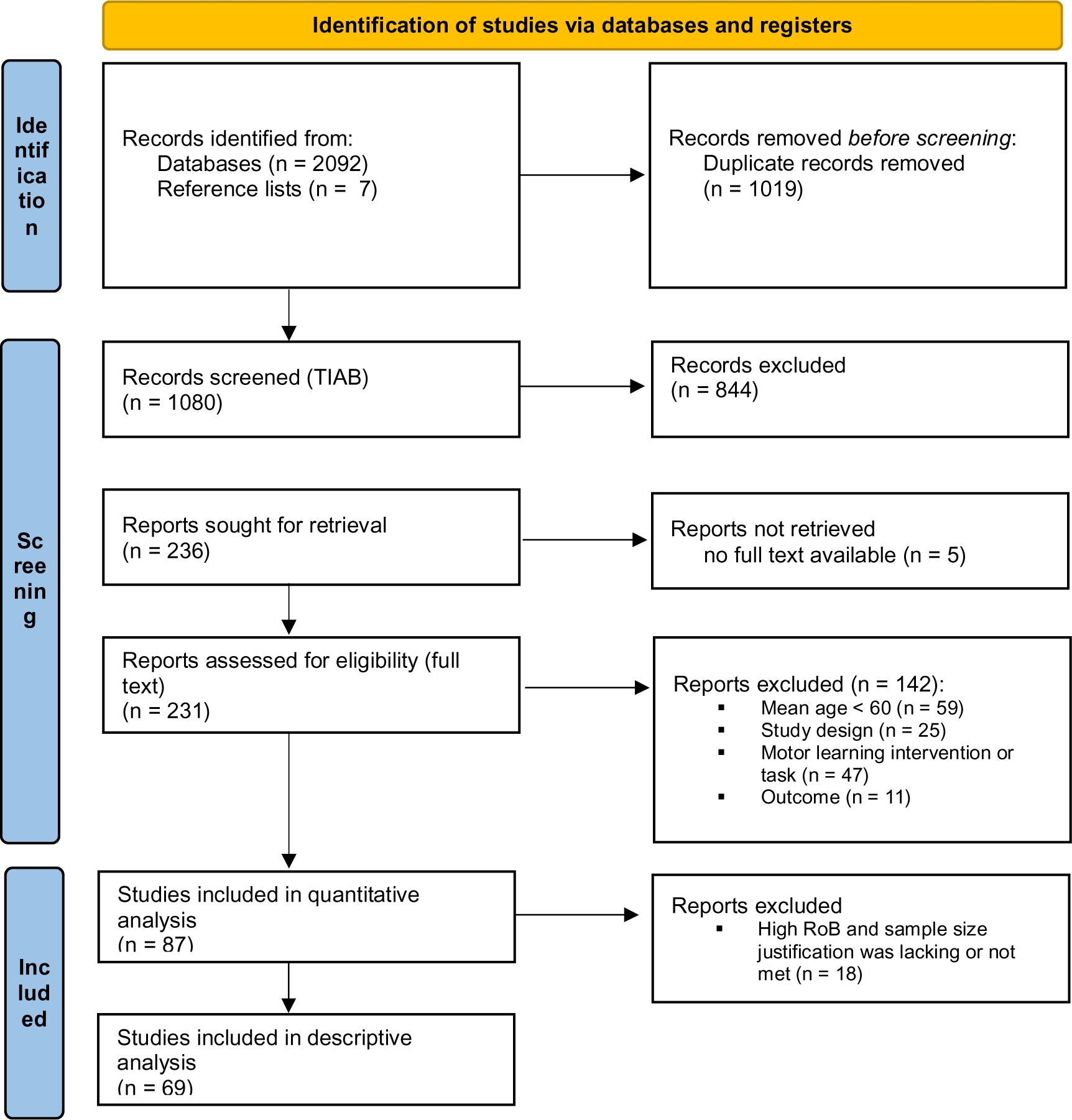
PRISMA flowchart for the inclusion of studies.

### Quantitative analysis (mapping)

In total, 87 studies were included. Six of the seven motor learning strategies were addressed (Figure 2). The most frequently described motor learning strategies were dual-task learning (n = 50 studies), mental practice (n = 19), and action observation (n = 12); no studies were found for discovery learning within these target populations. In total, eight different populations were identified within the included studies (Figure 2). Figure 3 visualises the number of studies published for each learning strategy over time.

**Figure 2.**
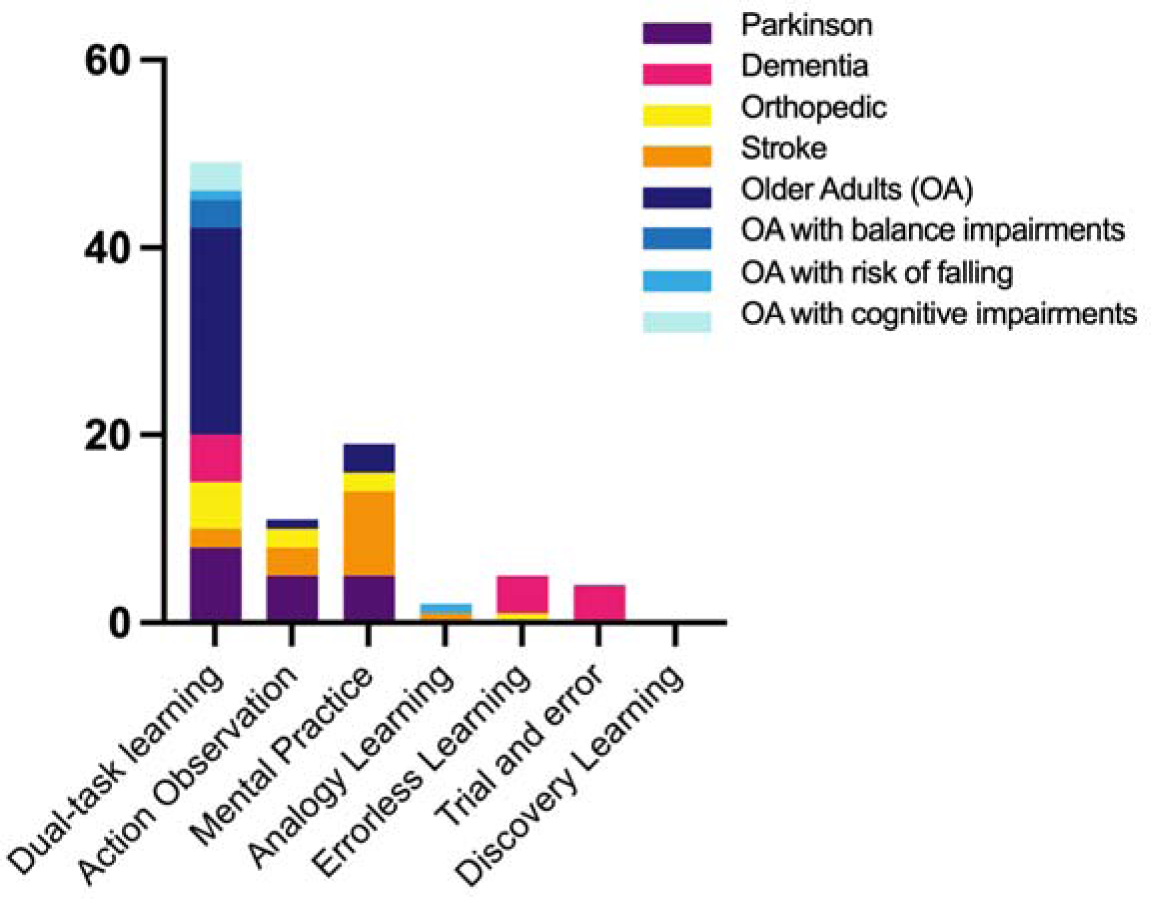
Visualisation of the number of studies identified per learning strategy including the sub-populations covered.

**Figure 3.**
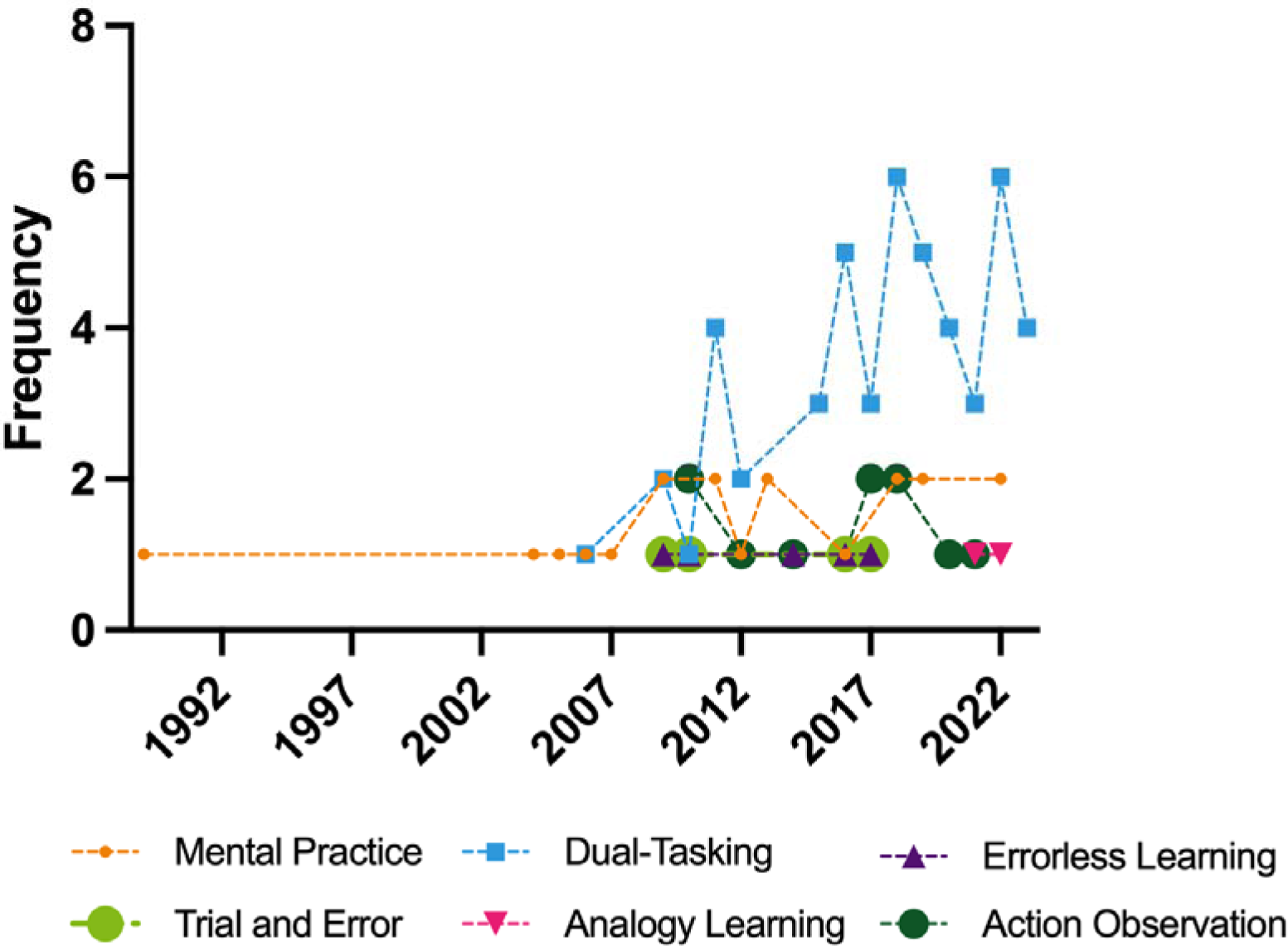
The year of publication per motor learning strategy. The identified motor learning strategies are based on the seven best-known and most-used motor learning strategies as embedded in the framework of Kleynen et al.^11^ Data of six motor learning strategies were included, as no studies for discovery learning were identified.

An overview of the risk of bias scores, power, group size, tasks, and between-group differences is reported in Table 2. Of the 87 studies, 5 scored well on both the RoB (low) and sample size justification, while 18 had a high RoB and did not report a sample size justification. Included studies’ group sizes ranged from 6 to 161 participants.

**Table 2.**
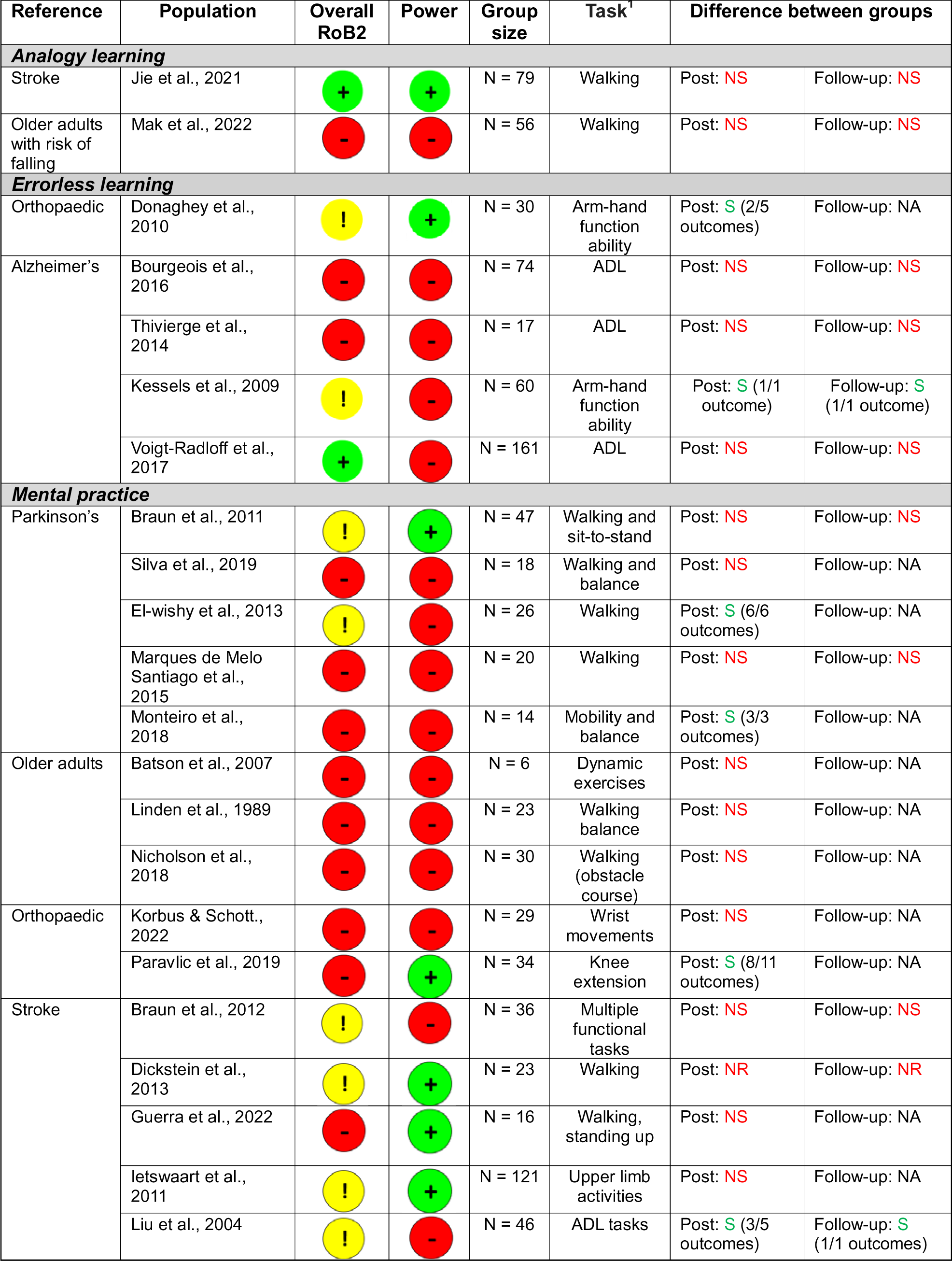
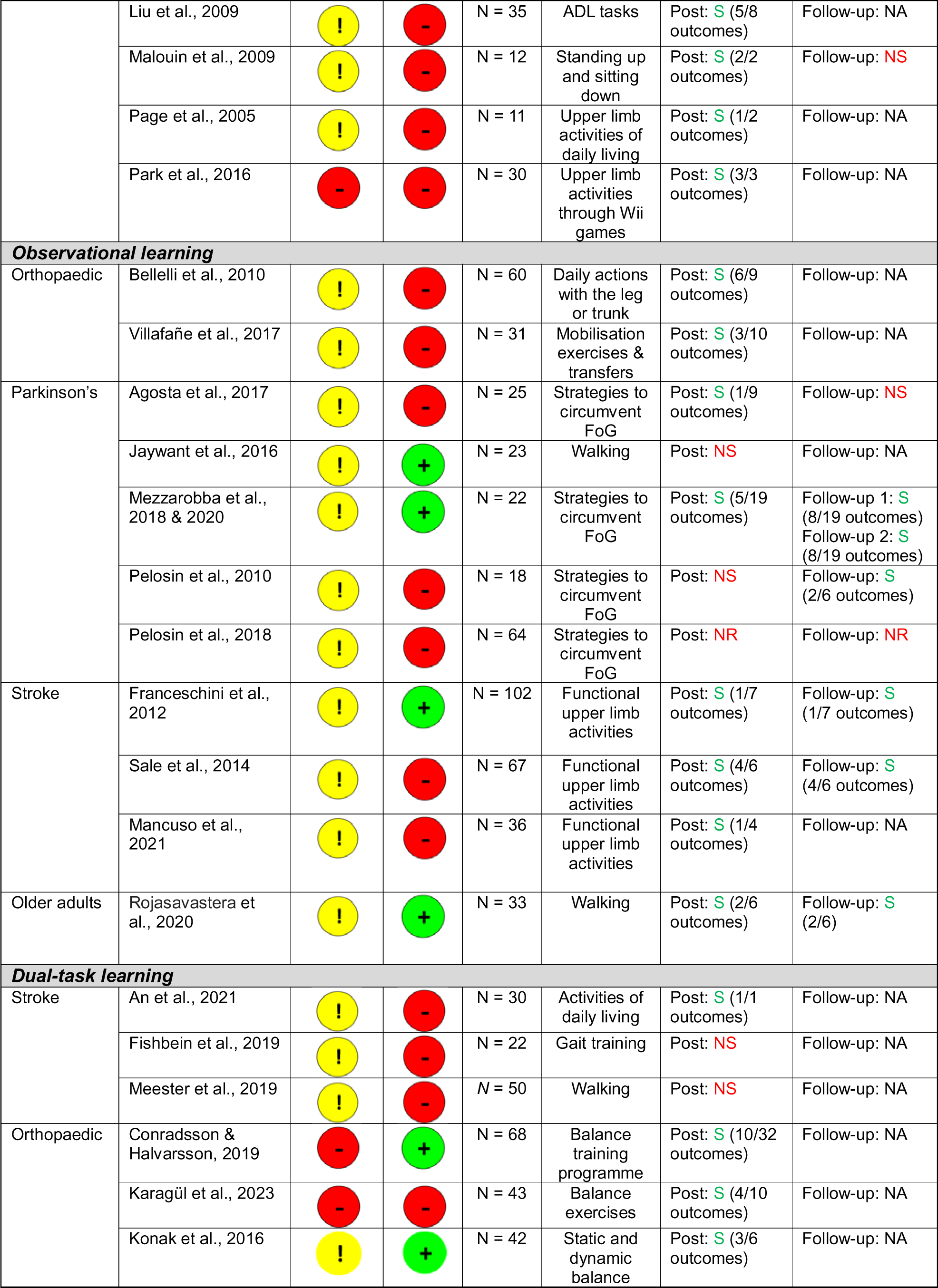
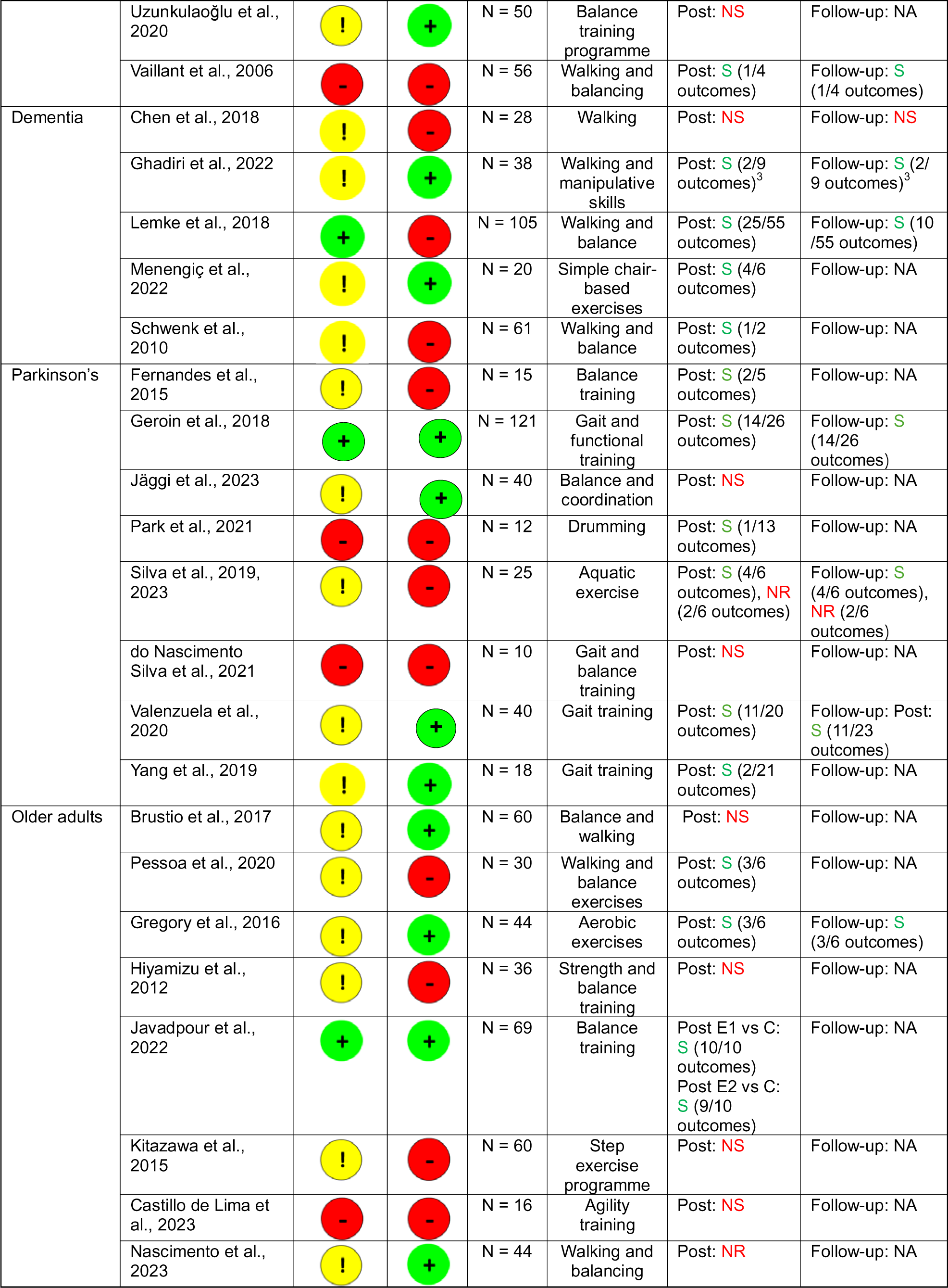
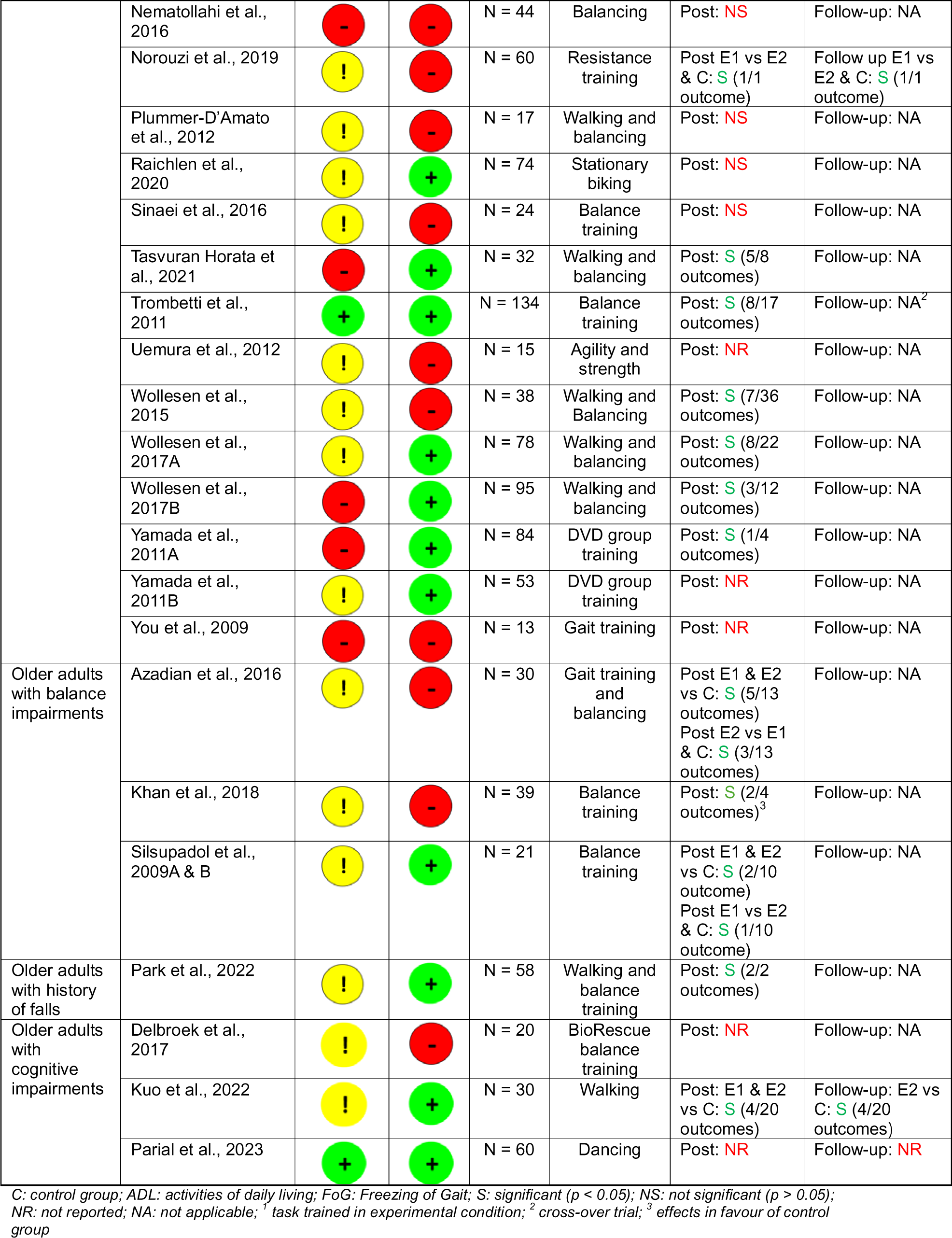
Risk of bias scores, power, group size, task, and overall outcome of the included studies

### Descriptive analysis

Sixty-nine studies were left for the descriptive analyses, the results which are presented below per learning strategy.

### Analogy learning

One study was included, with a sample size of 79 participants.^26^ The task trained in the experimental group was walking in community-dwelling individuals after stroke. The practised gait parameters were chosen based on the patients’ preferences and needs, as well as the clinical expertise of the therapists involved in the trial. The analogy learning instructions were personalised based on the individual’s walking impairment and preferences. The effectiveness of analogy learning was compared to an explicit motor learning intervention. No between-group differences were observed either post-intervention or at the follow-up (low RoB, appropriate sample size justification). The intervention’s duration was 3 weeks. The total intensity of training (i.e. the number of sessions multiplied by the duration of each session) was 270 minutes over 9 sessions. See Table 3 for more details.

**Table 3.** Detailed study characteristics of one analogy learning study

| **STROKE** |  |  |  |  |
| --- | --- | --- | --- | --- |
| **Reference** | **Population** | **Intervention** | **Amount of supervised practice** | **Measurement instruments, moments, and outcome** |
| Jie et al., 2021 | N = 79 persons after stroke in chronic phase | <u>Task trained in experimental group:</u><br>Walking. The to-be-improved gait parameter was chosen based on the therapist's expertise and the participants' needs. |  | <u>Measurement instrument:</u><br>- 10 Meter Walk Test<br>- Dual Task Costs motor task<br>- Dual Task Costs cognitive task<br>- Modified Dynamic Gait Index |
|  | <u>Analogy learning group:</u><br>n = 38; 64.6 yrs (SD: 9.4) | A learning environment was created where participants were not (or minimally) aware of the underlying rules of the practised motor skill. Participants received *personalised analogy instructions* based on their individual walking problems and preferences. | 9 sessions of 30 minutes in 3 weeks | <u>Measurement moment:</u><br>Pre- and post-intervention and 4-week follow-up<br><u>Between-group differences:</u><br>No significant differences in any of the outcomes |
|  | <u>Explicit learning group:</u><br>n = 41; 67.8 yrs (SD: 11.6) | A learning environment was created so participants were very aware of the learning process. Participants received *detailed explicit instructions* on how to improve their gait performance. | 9 sessions of 30 minutes in 3 weeks |  |

*N = number of participants in total, n = number of participants per group, SD = standard deviation, NA = not applicable, NR = not reported*

### Errorless learning and trial & error

Three studies were included, with sample sizes ranging from 30 to 161. Two studies included persons with Alzheimer’s, of which one study trained ADL activities that were based on the patients’ preferences and needs,^28^ and the other practised a functional arm-hand task from the Action Programme test.^29^ Furthermore, one other study included participants with transtibial amputations and examined in comparison to trial and error (n = 3).

The errorless learning intervention was structured in different ways. Frequent feed-forward instructions (i.e. ‘how to do’) were provided before initiation of the task. To minimise mistakes, some studies also provided cues verbally or pictorially (e.g. ^29,31^). In contrast to the errorless learning intervention, in the trial-and-error studies, participants were allowed to make mistakes and self-correct their performance. In some studies, open-ended questions about the task were posed to participants when repeated mistakes were observed (e.g. ^28^).

One study^29^ observed between-group differences in favour of the errorless learning intervention at the follow-up measurement; however, there are some concerns about RoB and a lack of sample size justification. Another study^30^ observed between-group differences post-intervention (some concerns about RoB, appropriate sample size justification). One study^28^ did not observe any between-group differences (low RoB, proposed sample size was not met in intention-to-treat analyses).

The total study duration of these interventions ranged from one single session^29,30^ up to 10 weeks.^28^ The total intensity of training (i.e. the number of sessions multiplied by the duration of each session) ranged from 15-30 minutes in one session^30^ to 540 minutes over 10 training sessions.^28^ Kessels and Olde Hensken^29^ did not specify the amount of time spent in each session. See Table 4 for more details.

**Table 4.** Detailed study characteristics of three studies on **errorless learning** and **trial and error**

| **ALZHEIMER'S** |  |  |  |  |
| --- | --- | --- | --- | --- |
| **Reference** | **Population** | **Intervention** | **Amount of supervised practice** | **Measurement instruments, moments, and outcome** |
| Kessels et al., 2009 | N = 60 persons with mild-to-moderate, severe dementia, or no dementia | <u>Task trained in experimental group:</u><br>Functional arm-hand tasks, e.g. removing a cork from a tube. |  | <u>Measurement instruments:</u><br>- Behavioural Assessment of the Dysexecutive Syndrome (BADS)<br><u>Measurement moments:</u><br>Pre- and post-intervention and 1-3 days follow-up<br><u>Between-group differences:</u><br>Difference in task performance on the BADS in favour of the errorless learning groups in persons with severe, mild-to-moderate, or no dementia. |
|  | <u>Errorless learning group:</u><br>Group 1 – severe dementia<br>*n* = 10; 83.6 yrs (SD: 8.1)<br>Group 2 – mild-to-moderate dementia<br>*n* = 10; 76.5 yrs (SD: 7.9)<br>Group 3 – controls: no dementia<br>*n* = 10; 72.7 yrs (SD: 11.0) | To prevent errors, participants received cues before completing a sequence of the to-be-completed task (e.g. 'You can use the hook to remove the lid'). | 1 session (duration of session NR) |  |
|  | <u>Trial-and-error learning group:</u><br>Group 1 – severe dementia<br>*n* = 10; 83.2 yrs (SD: 7.1)<br>Group 2 – mild-to-moderate dementia<br>*n* = 10; 77.1 yrs (SD: 9.4)<br>Group 3 – controls: no dementia<br>*n* = 10; 71.9 yrs (SD: 8.9) | Initially, cues were <u>not</u> provided to complete the task. Cues were only provided (in the second instance) if they were unable to find and complete the correct next step, i.e. errors were made. | 1 session (duration of session NR) |  |
| Voigt-Radloff et al., 2017 | N = 161 persons with Alzheimer's or mixed-type dementia | <u>Task trained in experimental group:</u> By shared decision-making, participants practised two tasks relevant to daily living. |  | <u>Measurement instruments:</u><br>Core Elements Method (CEM: a 7-point scale for each task where 1 = not performed at all as trained by the therapist; 7 = performed exactly as trained by the therapist)<br><u>Measurement moments:</u><br>Pre- and post-intervention, and 6- and 16-week follow-up<br><u>Between-group differences:</u><br>No significant differences in any of the outcomes. |
|  | <u>Errorless learning group:</u><br>*n* = 81; 77.1 yrs (SD: 7.8) | The therapist divided the task into appropriate steps, demonstrated and (verbally) guided the participants through the steps. If potential errors occurred, then the therapist anticipated through providing a short demonstration of the correct performance, i.e. errors were prevented | 9 sessions of 60 min in 10 weeks |  |
|  | <u>Trial-and-error group:</u><br>*n* = 80; 76.1 yrs (SD: 6.8) | Participants were asked to perform the task but did not receive instructions or demonstration. After 3 insufficient trials, open-ended questions were asked about the purpose of the task to find solutions. | 9 sessions of 60 min in 10 weeks |  |

| **ORTHOPAEDICS** |  |  |  |  |
| --- | --- | --- | --- | --- |
| **Reference** | **Population** | **Intervention** | **Amount of supervised practice** | **Measurement instruments, moments, and outcome** |
| Donaghey et al., 2010 | N = 30 persons with unilateral transtibial amputations | <u>Task trained in experimental group:</u><br>Fitting a prosthetic limb. | All participants were first shown how to put on the prosthetic limb. | <u>Measurement instruments:</u><br>Video analyses of the final trial included:<br>- Total number of correct steps<br>- Number of omissions<br>- Number of deviations<br>- Number of hesitations<br>- Time taken to complete the fitting sequence<br><u>Measurement moments:</u><br>Post-intervention<br><u>Between-group differences:</u><br>Difference in correct steps in the fitting sequence and number of errors in favour of the errorless learning group |
|  | <u>Errorless learning group:</u><br>n = 15; 62 yrs (SD: 14.6) | The participants were 'talked through' the sequence of tasks, such that they were unable to make any errors. Participants received the appropriate limb parts needed to move correctly to the next stage of the fitting sequence. | 1 session of 15-30 min |  |
|  | <u>Trial-and-error group:</u><br>n = 15; 66 yrs (SD: 6.8) | Learning proceeded via a trial-and-error process. | 1 session of 15-30 min |  |

N = number of participants in total, n = number of participants per group, SD = standard deviation, NA = not applicable, NR = not reported, BADS = Behavioural Assessment of the Dysexecutive Syndrome,

### Mental practice

Eleven studies were included with sample sizes ranging from 11 to 121 participants. Two studies included people with Parkinson’s, both focusing on gait.^33,34^ One study included participants after total knee arthroplasty and practised knee extension.^35^ Eight studies included people after stroke, two of which focusing on gait,^36,37^ three on upper limb activities,^38–40^ two on daily life activities,^41–43^ and one relaxation (n = 2), care as usual (n = 5), cognitive exercises (or mental rehearsal; n = 3), and standardised activities for the upper limbs (n = 1), and two studies specifically described that the intervention included a demonstration-then-practice element.

Mental practice interventions were often based on standardised protocols, scripts, or frameworks. The different stages of the mental practice intervention often included familiarisation with the task (e.g. analysis of the task sequence) and mental practice aspects (e.g. kinematic components), followed by internal imagery, mental rehearsal, and overt task performance. Two studies used audio instructions, while in one other study, the mental practice intervention was guided by a computer program. Different types of mental practice reported in the studies included kinaesthetic, visual, and motivational imagery.

One study^42^ observed between-group differences in favour of the intervention post-intervention and at the follow-up measurement (some concerns about RoB, sample size justification lacking). Five studies^34,35,39,43,44^ observed between-group differences in favour of the mental practice intervention post-intervention (RoB ranged from some concerns to high, one with appropriate sample size justification). Three studies^33,38,41^ did not find any between-group differences (all with some concerns about RoB, two with appropriate sample size justification), and one study^36^ did not calculate any between-group differences.

The total study duration of these interventions ranged from 3 weeks^36^ ^42,43^ up to 6 weeks.^39^ The total intensity of training (i.e. the number of sessions multiplied by the duration of each session) ranged from 180 minutes over 12 sessions^36^ to 900 minutes over 15 training sessions.^42,43^ Braun et al.^33,41^ did not specify the amount of time spent in each session. See Table 5 for more details.

**Table 5.**
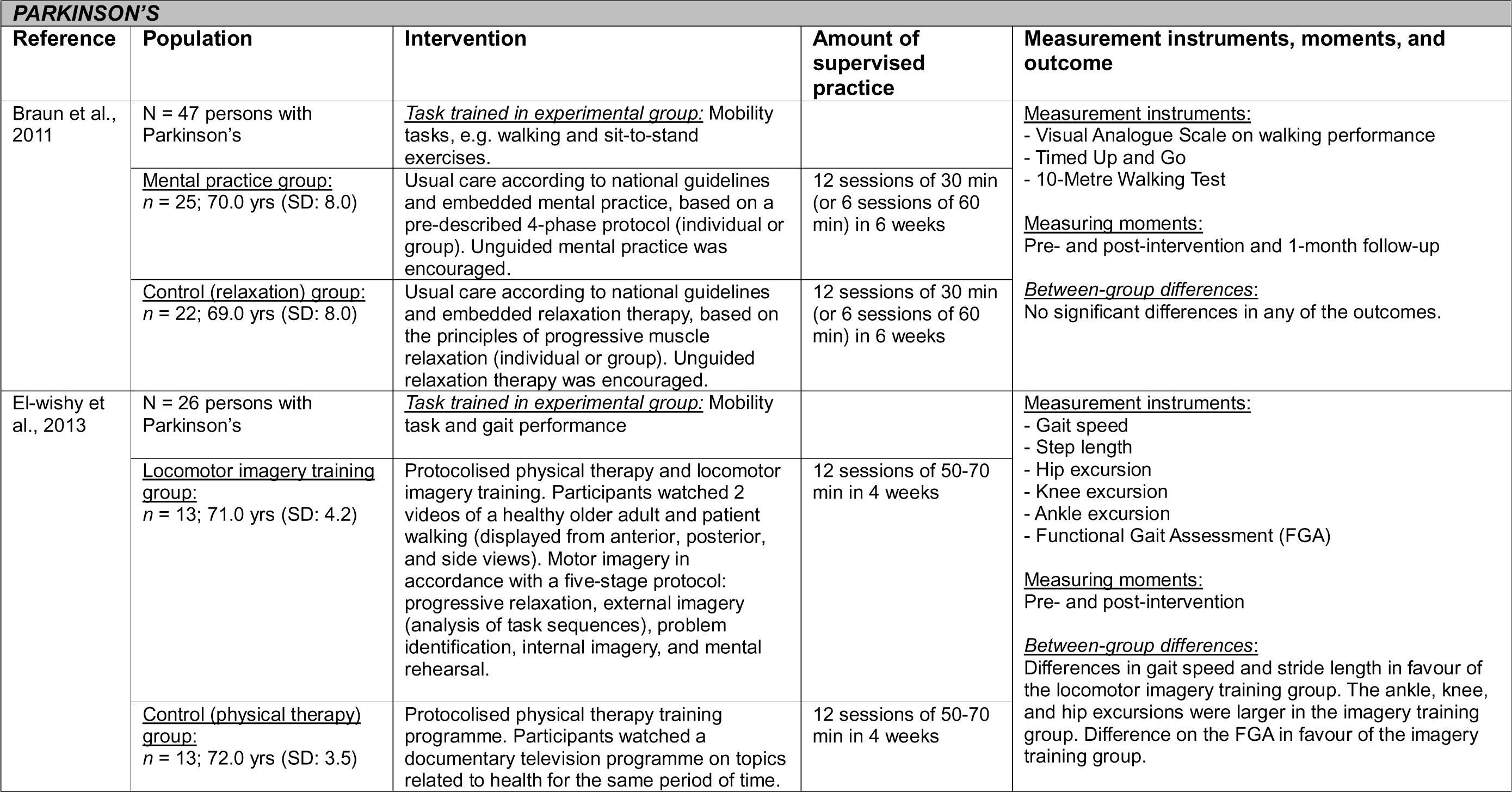
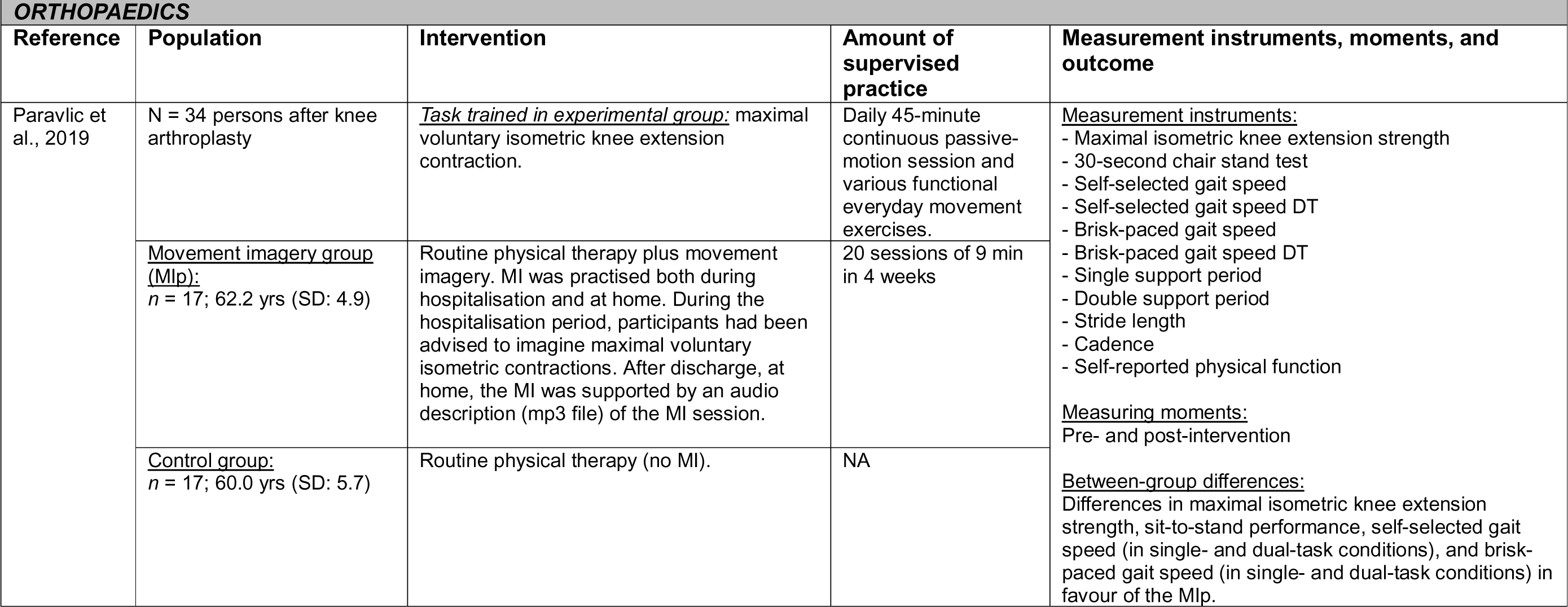
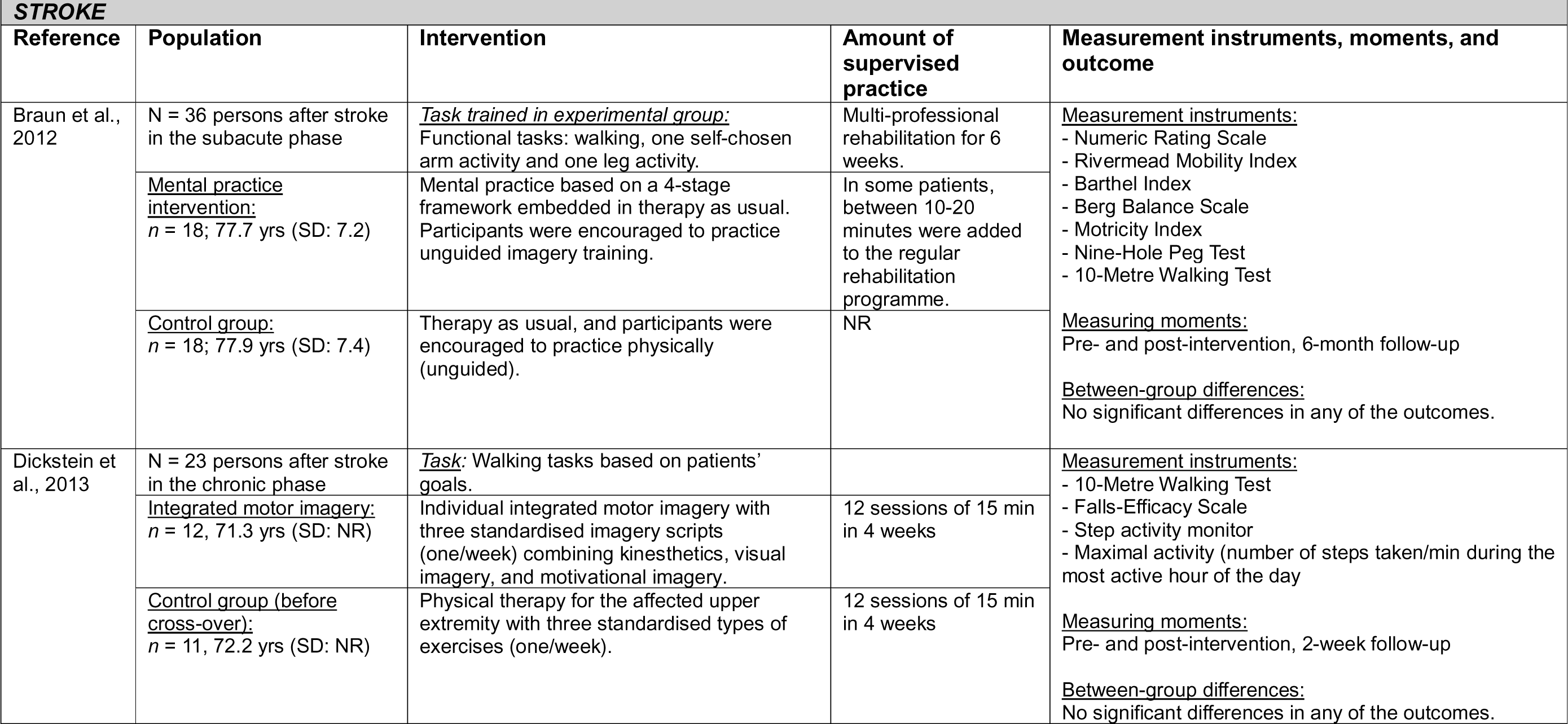
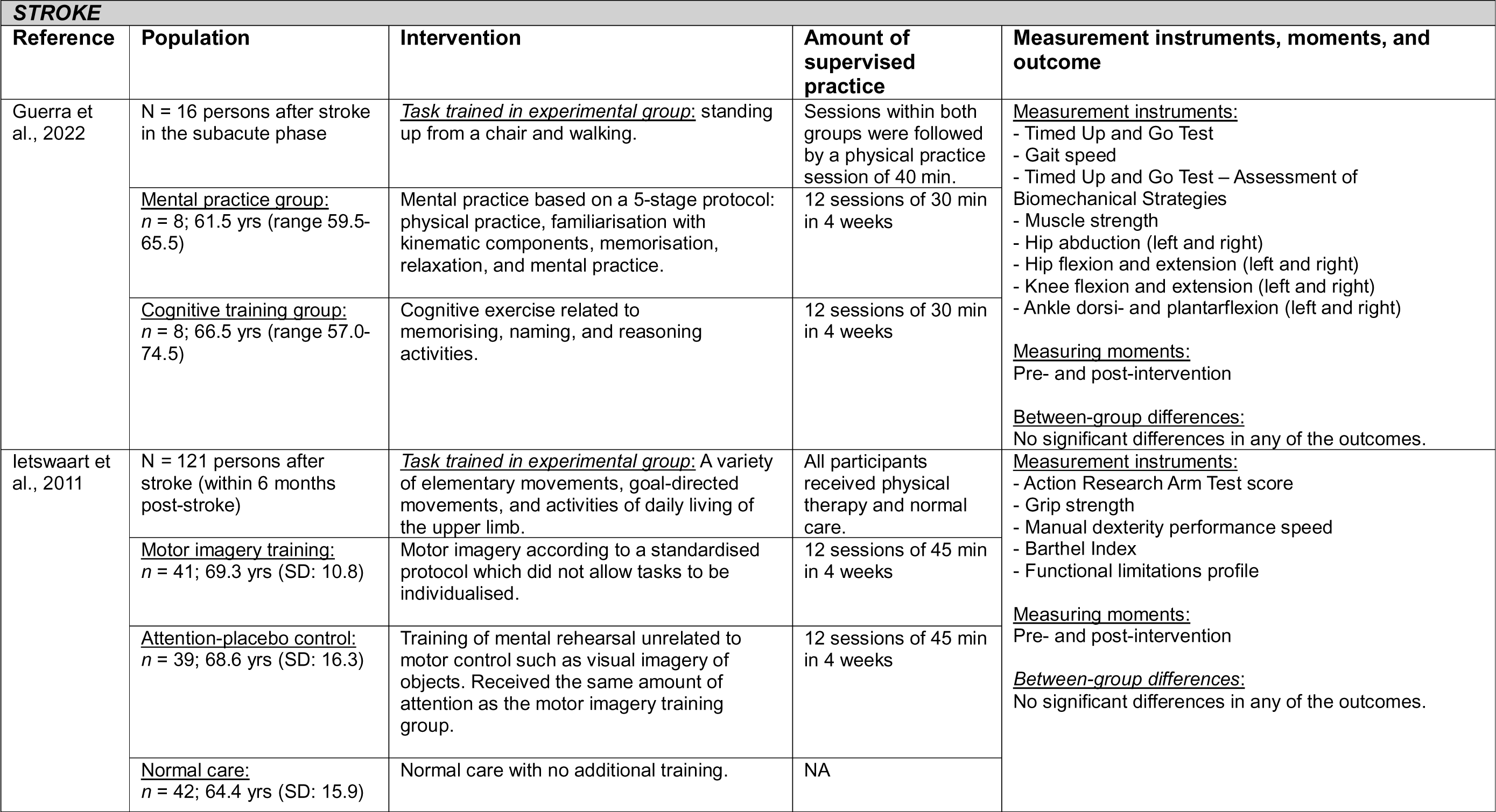
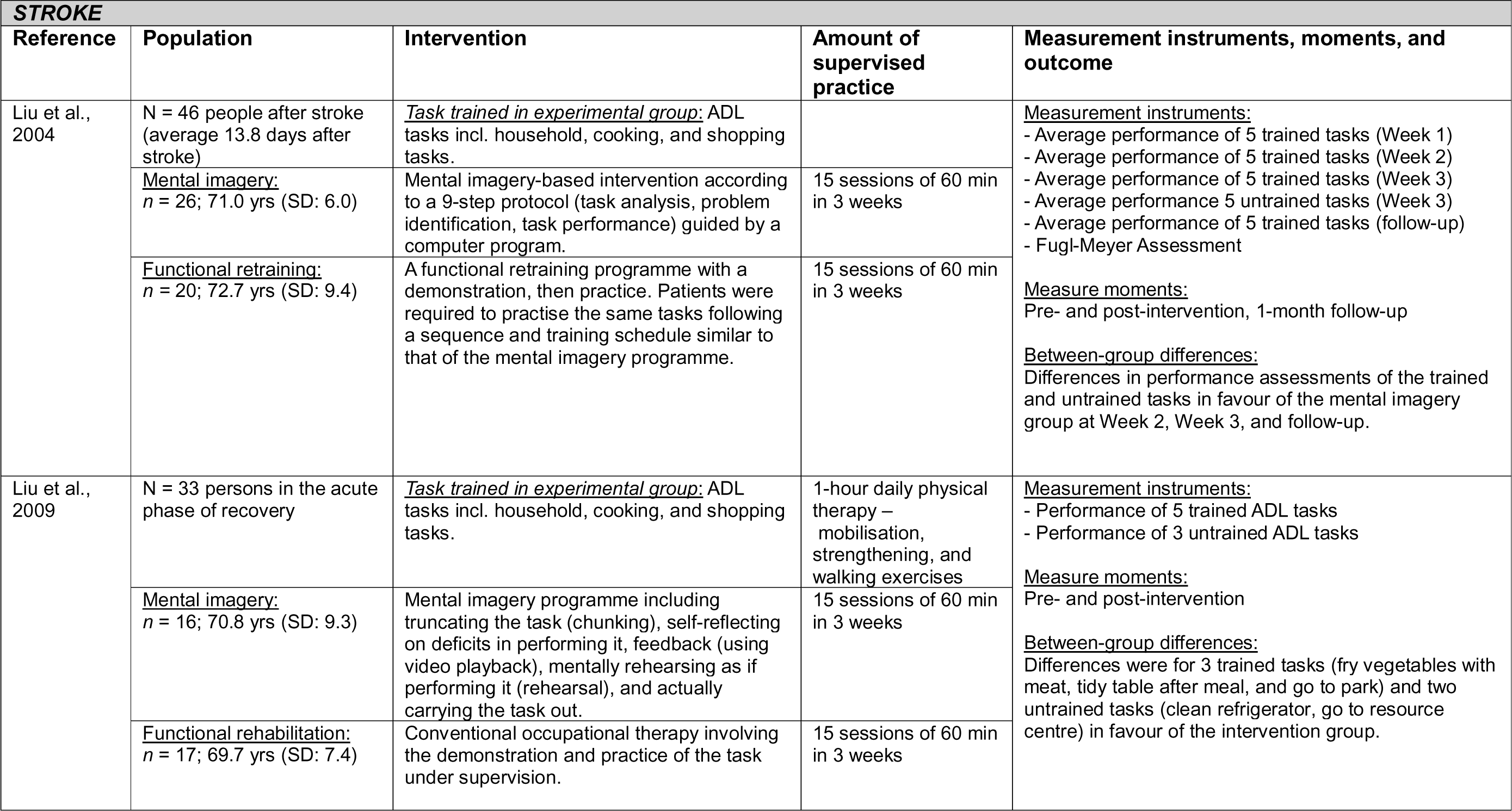
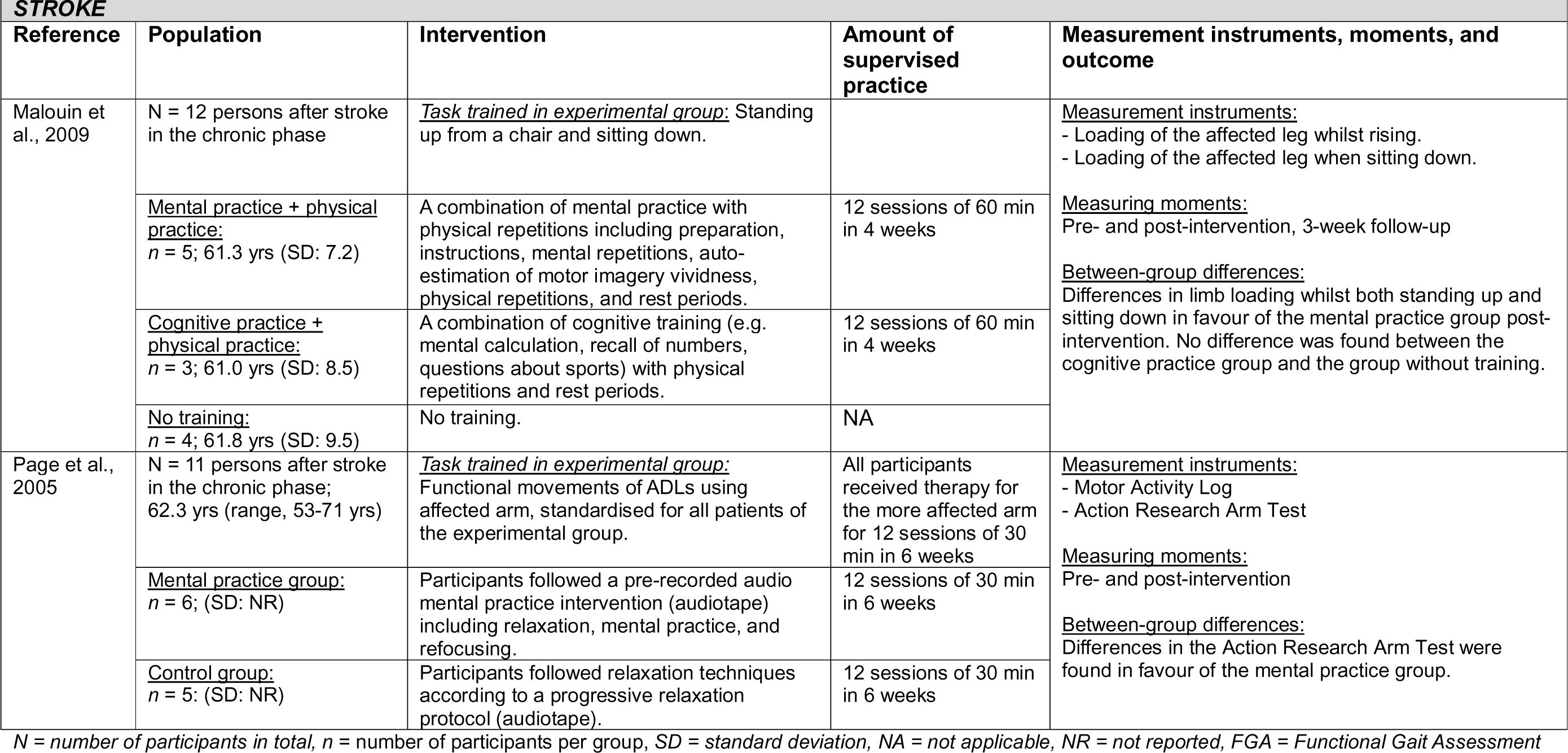
Detailed study characteristics of 11 studies on **mental practice**

### Observational learning

Eleven included studies had sample sizes ranging from 18 to 102 participants. Three studies included people after stroke who only practised upper limb activities.^51–53^ Five studies included people with Parkinson’s, focusing on gait in general (n = 1)^54^ or freezing of gait in particular (n = 4).^25,55–57^ Two studies included orthopaedic patients in which daily activities,^58^ mobilisation exercises, and transfers^59^ were trained. One study included older adults who practised walking.^60^

In all studies, observational learning was combined with or integrated into different types of functional training. In all studies, observational learning was applied through watching short movies of the task, exercise, or strategy to be learned. One study used videos that were composed of images and sounds (sonification).^25^ Most studies (n = 9) investigated the effects of observing functional movements in comparison to observation of landscape videos or abstract pictures.^51,53–60^ Two studies compared observational learning to a functional training intervention without (action) observation.^52,61^

Four studies^51,53,60,61^ observed between-group differences in favour of the action observation intervention post-intervention and at the follow-up measurement (all with some concerns about RoB, three with appropriate sample size justification). Five studies^52,55,56,58,59^ observed between-group differences in favour of the intervention at either of the measurement points (all with some concerns about RoB, two with appropriate sample size justification). One study^54^ did not find any between-group differences (some concerns about RoB, appropriate sample size justification), and one study^57^ did not calculate any between-group differences.

The total study duration of these interventions ranged from 8 days^54^ up to 8 weeks.^61^ The total intensity of training (i.e. the number of sessions multiplied by the duration of each session) ranged from 432 minutes over 18 sessions^58^ to 960 minutes over 16 training sessions.^61^ Jaywant et al.^54^ did not specify the amount of time spent in each session. See Table 6 for more details.

**Table 6.**
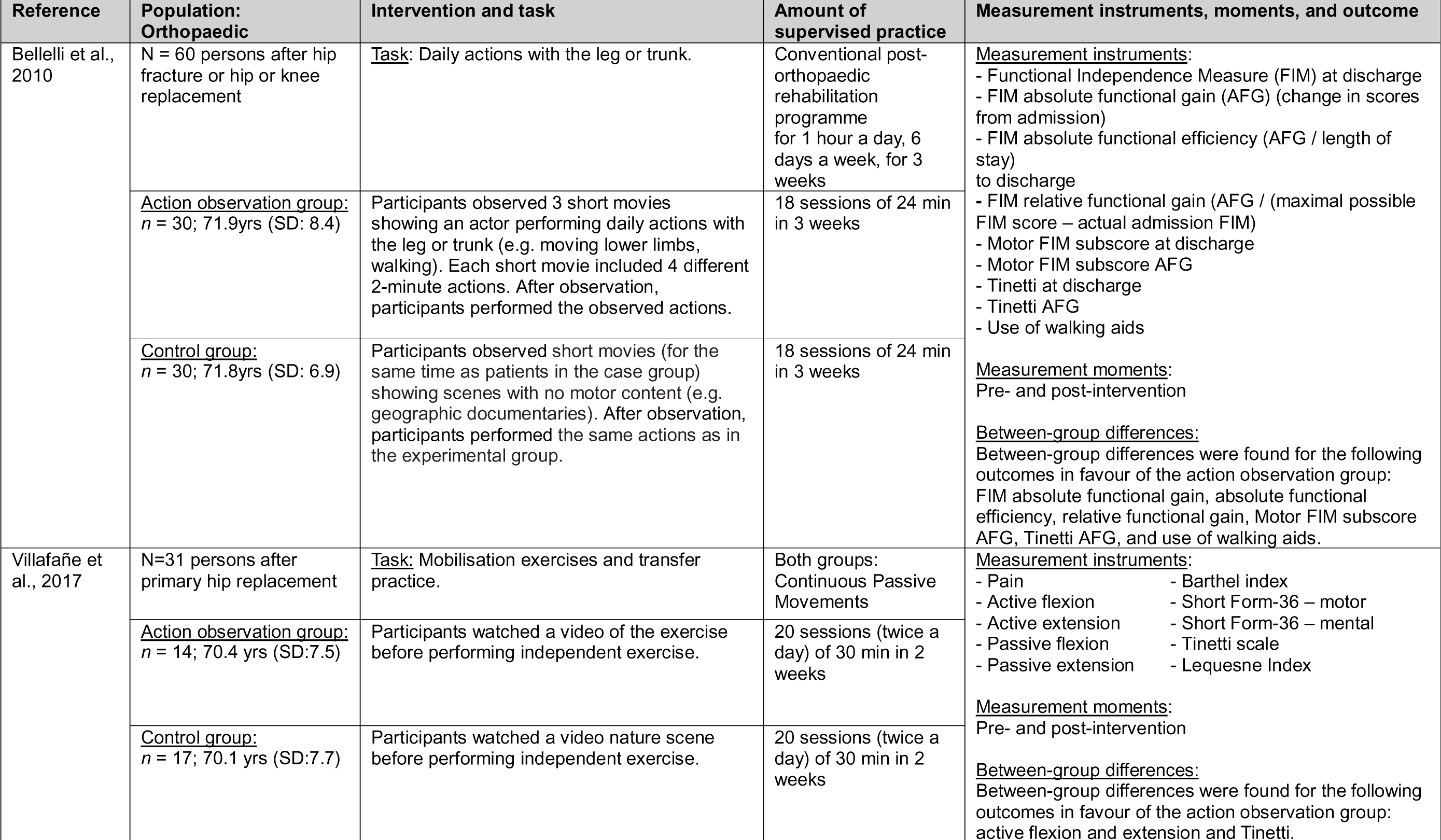
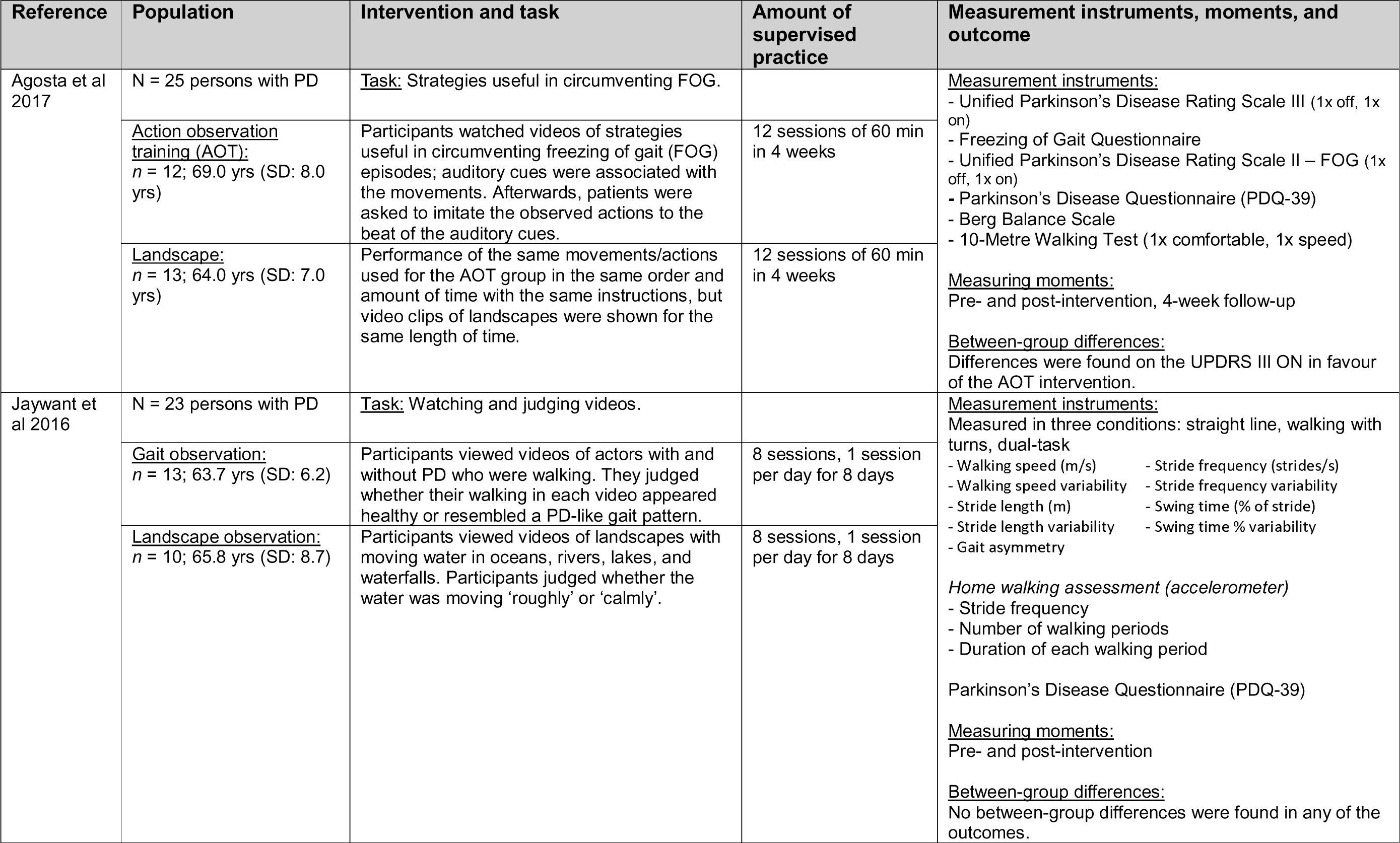
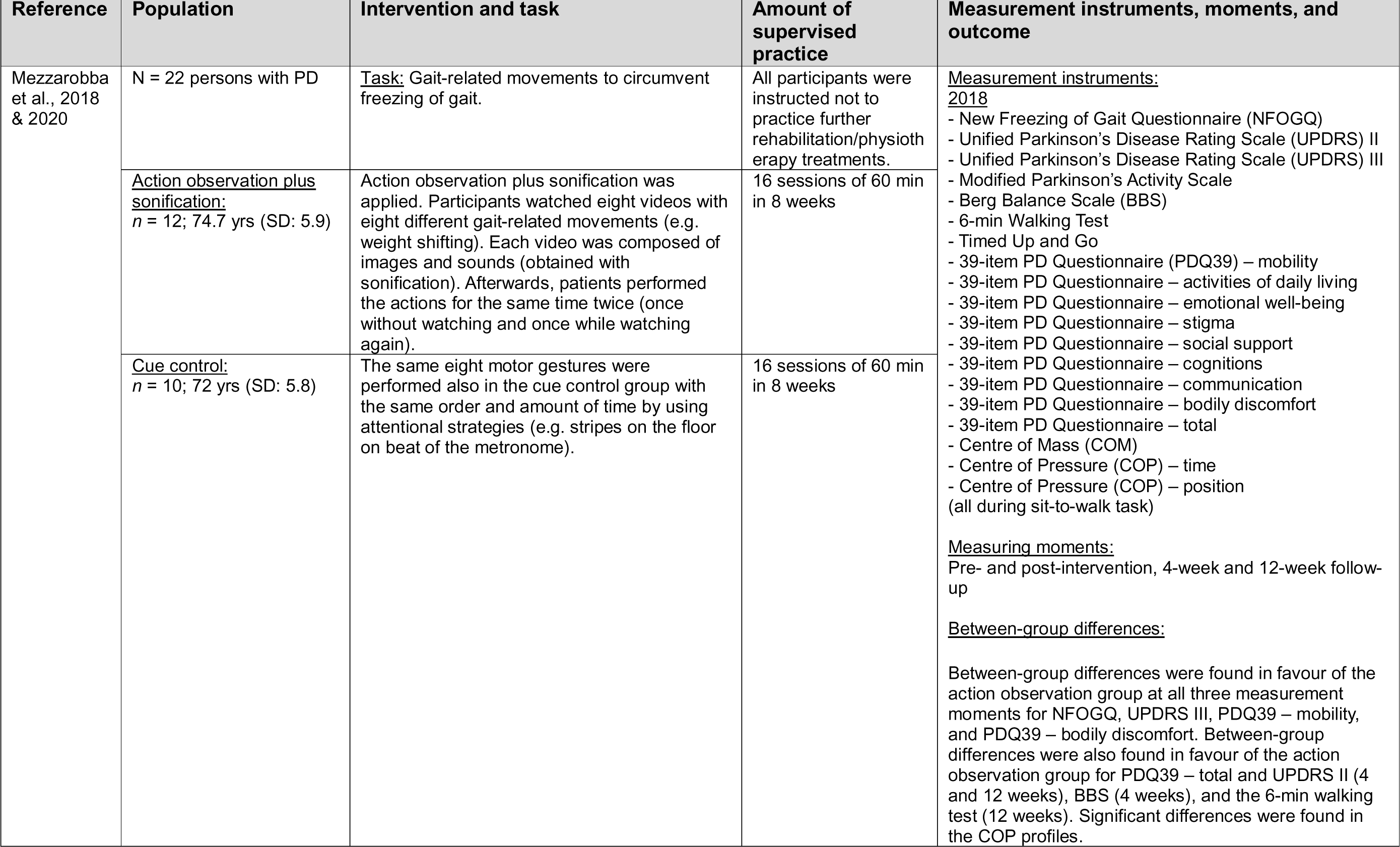
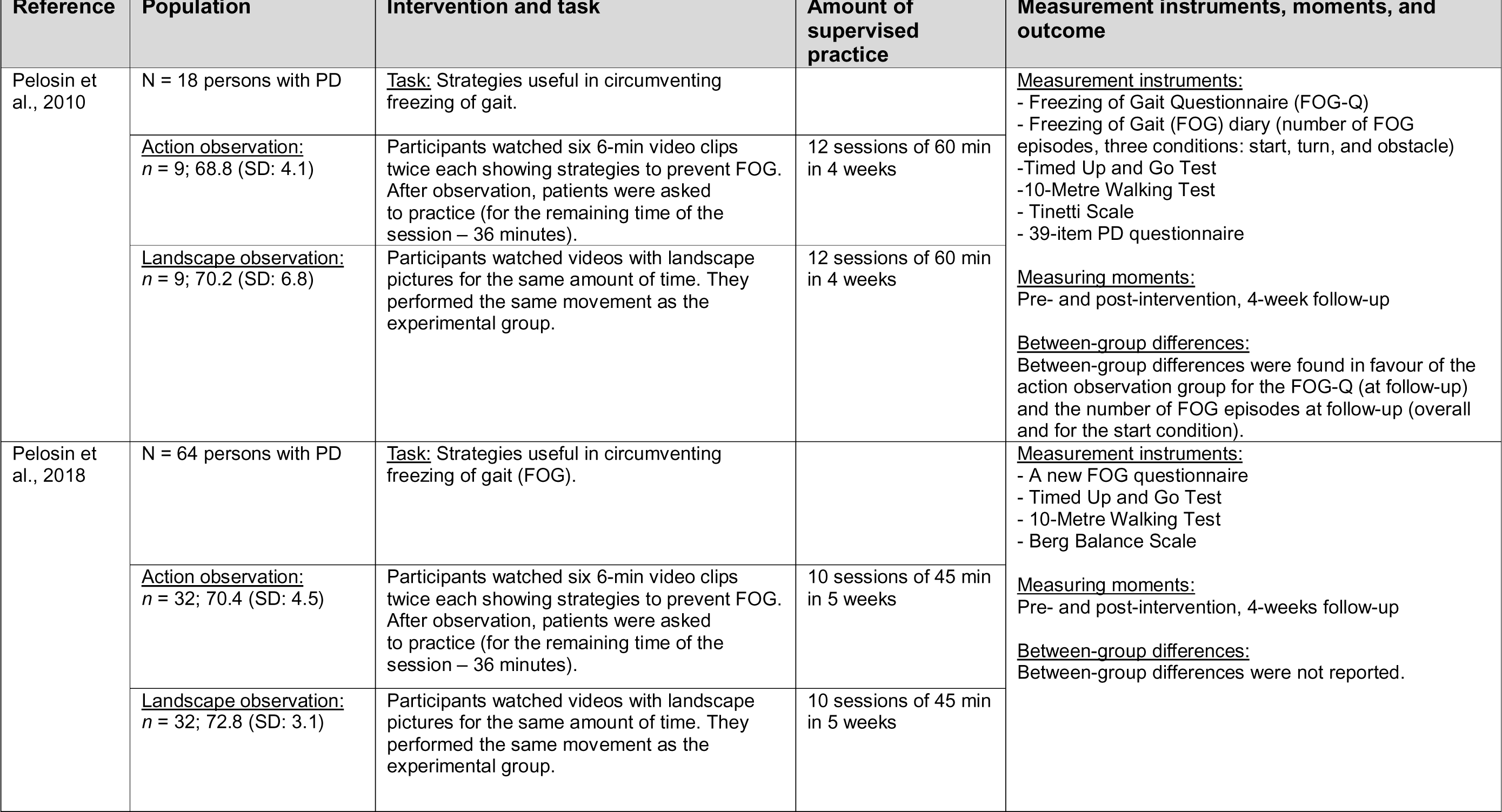
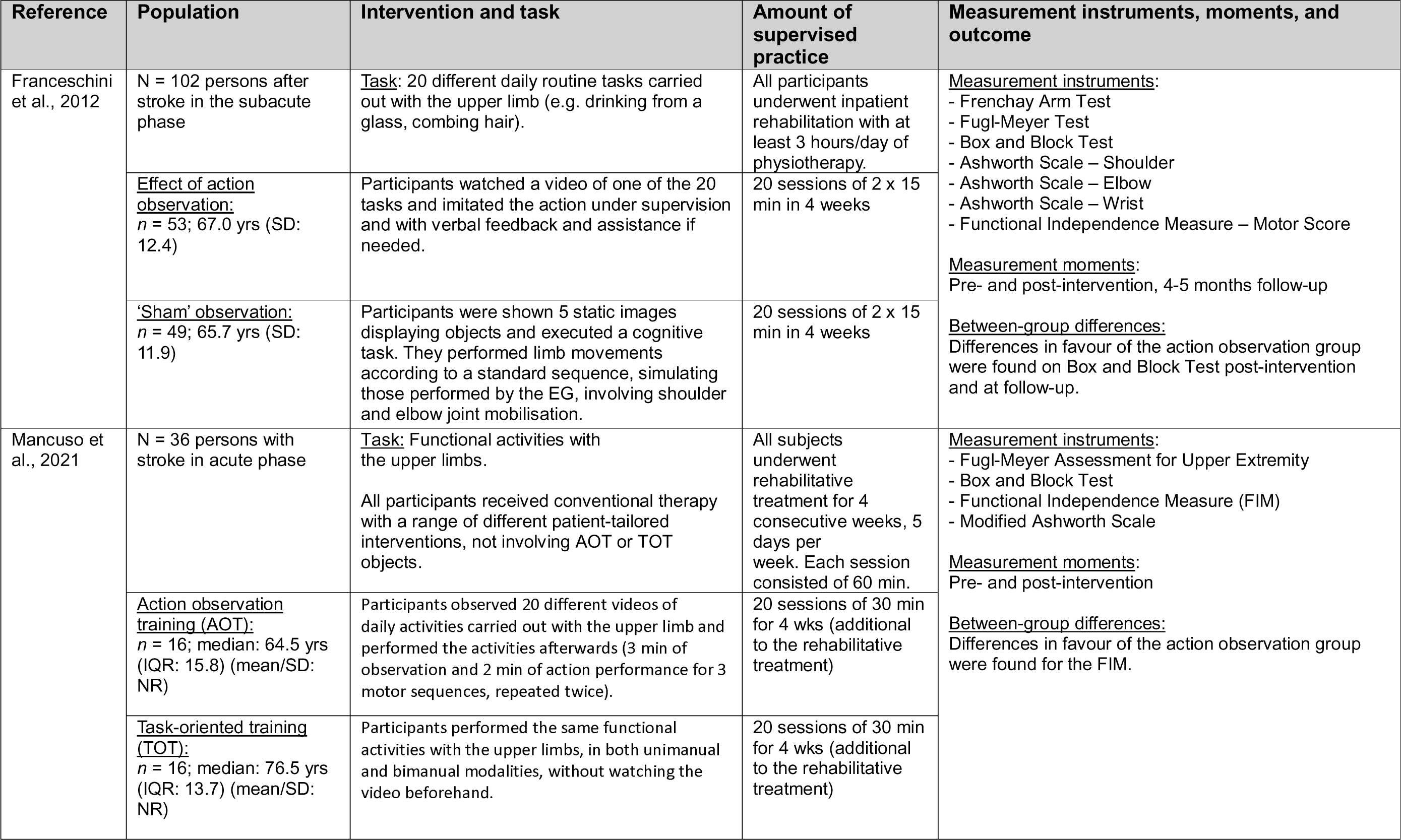
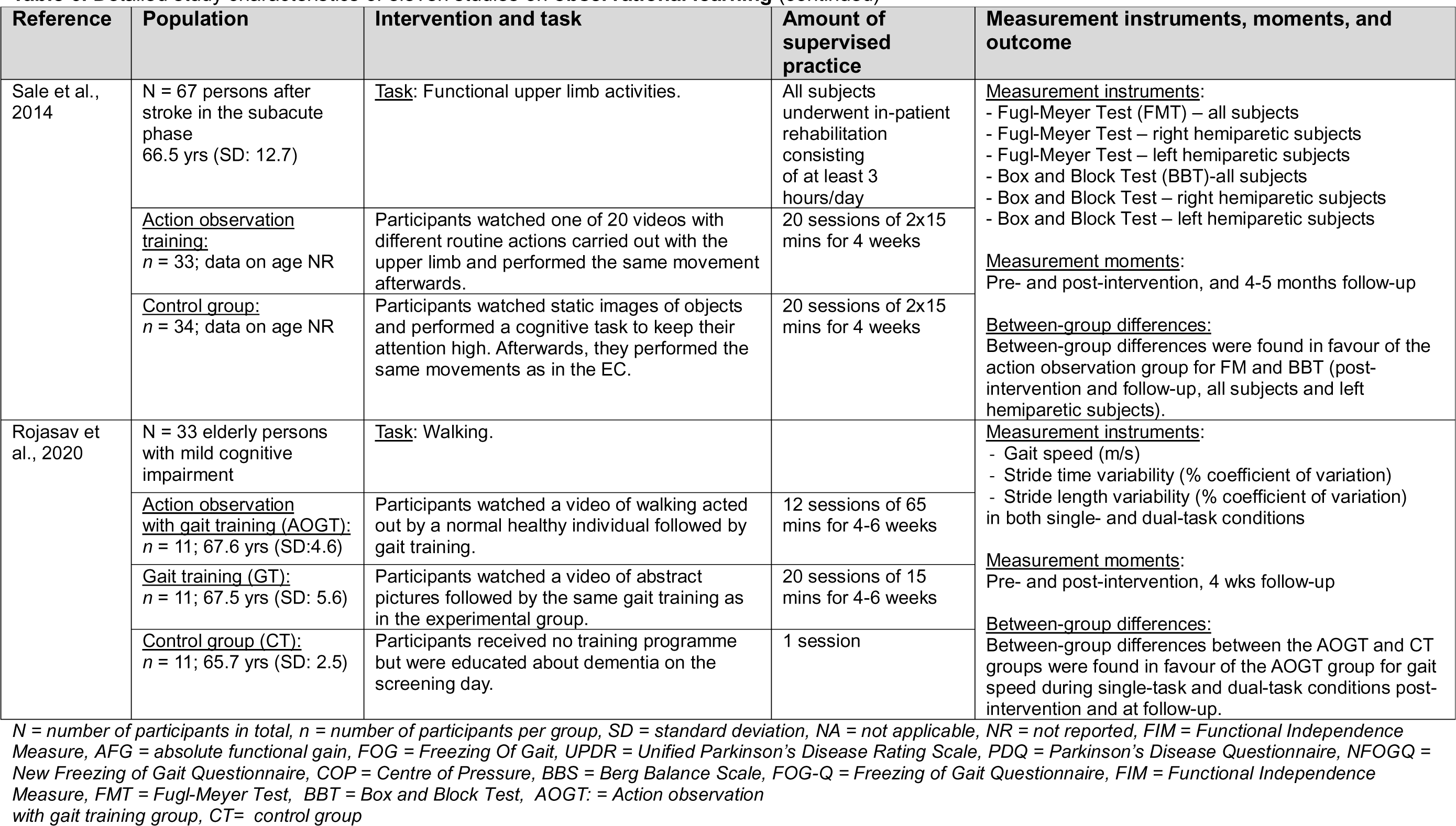
Detailed study characteristics of eleven studies on **observational learning**

| Reference | Population:<br>Orthopaedic | Intervention and task | Amount of<br>supervised practice | Measurement instruments, moments, and outcome |
| --- | --- | --- | --- | --- |
| Bellelli et al., 2010 | N = 60 persons after hip fracture or hip or knee replacement | <u>Task:</u> Daily actions with the leg or trunk. | Conventional post-orthopaedic rehabilitation programme for 1 hour a day, 6 days a week, for 3 weeks | <u>Measurement instruments:</u><br>- Functional Independence Measure (FIM) at discharge<br>- FIM absolute functional gain (AFG) (change in scores from admission)<br>- FIM absolute functional efficiency (AFG / length of stay) to discharge<br>- FIM relative functional gain (AFG / (maximal possible FIM score – actual admission FIM))<br>- Motor FIM subscore at discharge<br>- Motor FIM subscore AFG<br>- Tinetti at discharge<br>- Tinetti AFG<br>- Use of walking aids<br><u>Measurement moments:</u><br>Pre- and post-intervention<br><u>Between-group differences:</u><br>Between-group differences were found for the following outcomes in favour of the action observation group: FIM absolute functional gain, absolute functional efficiency, relative functional gain, Motor FIM subscore AFG, Tinetti AFG, and use of walking aids. |
|  | <u>Action observation group:</u><br>n = 30; 71.9yrs (SD: 8.4) | Participants observed 3 short movies showing an actor performing daily actions with the leg or trunk (e.g. moving lower limbs, walking). Each short movie included 4 different 2-minute actions. After observation, participants performed the observed actions. | 18 sessions of 24 min in 3 weeks |  |
|  | <u>Control group:</u><br>n = 30; 71.8yrs (SD: 6.9) | Participants observed short movies (for the same time as patients in the case group) showing scenes with no motor content (e.g. geographic documentaries). After observation, participants performed the same actions as in the experimental group. | 18 sessions of 24 min in 3 weeks |  |
| Villafañe et al., 2017 | N=31 persons after primary hip replacement | <u>Task:</u> Mobilisation exercises and transfer practice. | Both groups:<br>Continuous Passive Movements | <u>Measurement instruments:</u><br>- Pain<br>- Active flexion<br>- Active extension<br>- Passive flexion<br>- Passive extension<br>- Barthel index<br>- Short Form-36 – motor<br>- Short Form-36 – mental<br>- Tinetti scale<br>- Lequesne Index<br><u>Measurement moments:</u><br>Pre- and post-intervention<br><u>Between-group differences:</u><br>Between-group differences were found for the following outcomes in favour of the action observation group: active flexion and extension and Tinetti. |
|  | <u>Action observation group:</u><br>n = 14; 70.4 yrs (SD:7.5) | Participants watched a video of the exercise before performing independent exercise. | 20 sessions (twice a day) of 30 min in 2 weeks |  |
|  | <u>Control group:</u><br>n = 17; 70.1 yrs (SD:7.7) | Participants watched a video nature scene before performing independent exercise. | 20 sessions (twice a day) of 30 min in 2 weeks |  |

| Reference | Population | Intervention and task | Amount of supervised practice | Measurement instruments, moments, and outcome |
| --- | --- | --- | --- | --- |
| Agosta et al 2017 | N = 25 persons with PD | <u>Task</u>: Strategies useful in circumventing FOG. |  | <u>Measurement instruments</u>:<br>- Unified Parkinson's Disease Rating Scale III (1x off, 1x on)<br>- Freezing of Gait Questionnaire<br>- Unified Parkinson's Disease Rating Scale II – FOG (1x off, 1x on)<br>- Parkinson's Disease Questionnaire (PDQ-39)<br>- Berg Balance Scale<br>- 10-Metre Walking Test (1x comfortable, 1x speed)<br><u>Measuring moments</u>:<br>Pre- and post-intervention, 4-week follow-up<br><u>Between-group differences</u>:<br>Differences were found on the UPDRS III ON in favour of the AOT intervention. |
|  | <u>Action observation training (AOT)</u>:<br>n = 12; 69.0 yrs (SD: 8.0 yrs) | Participants watched videos of strategies useful in circumventing freezing of gait (FOG) episodes; auditory cues were associated with the movements. Afterwards, patients were asked to imitate the observed actions to the beat of the auditory cues. | 12 sessions of 60 min in 4 weeks |  |
|  | <u>Landscape</u>:<br>n = 13; 64.0 yrs (SD: 7.0 yrs) | Performance of the same movements/actions used for the AOT group in the same order and amount of time with the same instructions, but video clips of landscapes were shown for the same length of time. | 12 sessions of 60 min in 4 weeks |  |
| Jaywant et al 2016 | N = 23 persons with PD | <u>Task</u>: Watching and judging videos. |  | <u>Measurement instruments</u>:<br>Measured in three conditions: straight line, walking with turns, dual-task<br>- Walking speed (m/s) - Stride frequency (strides/s)<br>- Walking speed variability - Stride frequency variability<br>- Stride length (m) - Swing time (% of stride)<br>- Stride length variability - Swing time % variability<br>- Gait asymmetry<br>*Home walking assessment (accelerometer)*<br>- Stride frequency<br>- Number of walking periods<br>- Duration of each walking period<br>Parkinson's Disease Questionnaire (PDQ-39)<br><u>Measuring moments</u>:<br>Pre- and post-intervention<br><u>Between-group differences</u>:<br>No between-group differences were found in any of the outcomes. |
|  | <u>Gait observation</u>:<br>n = 13; 63.7 yrs (SD: 6.2) | Participants viewed videos of actors with and without PD who were walking. They judged whether their walking in each video appeared healthy or resembled a PD-like gait pattern. | 8 sessions, 1 session per day for 8 days |  |
|  | <u>Landscape observation</u>:<br>n = 10; 65.8 yrs (SD: 8.7) | Participants viewed videos of landscapes with moving water in oceans, rivers, lakes, and waterfalls. Participants judged whether the water was moving 'roughly' or 'calmly'. | 8 sessions, 1 session per day for 8 days |  |

| Reference | Population | Intervention and task | Amount of supervised practice | Measurement instruments, moments, and outcome |
| --- | --- | --- | --- | --- |
| Mezzarobba et al., 2018 & 2020 | N = 22 persons with PD | <u>Task:</u> Gait-related movements to circumvent freezing of gait. | All participants were instructed not to practice further rehabilitation/physiotherapy treatments. | <u>Measurement instruments:</u><br>**2018**<br>- New Freezing of Gait Questionnaire (NFOGQ)<br>- Unified Parkinson's Disease Rating Scale (UPDRS) II<br>- Unified Parkinson's Disease Rating Scale (UPDRS) III<br>- Modified Parkinson's Activity Scale<br>- Berg Balance Scale (BBS)<br>- 6-min Walking Test<br>- Timed Up and Go<br>- 39-item PD Questionnaire (PDQ39) – mobility<br>- 39-item PD Questionnaire – activities of daily living<br>- 39-item PD Questionnaire – emotional well-being<br>- 39-item PD Questionnaire – stigma<br>- 39-item PD Questionnaire – social support<br>- 39-item PD Questionnaire – cognitions<br>- 39-item PD Questionnaire – communication<br>- 39-item PD Questionnaire – bodily discomfort<br>- 39-item PD Questionnaire – total<br>- Centre of Mass (COM)<br>- Centre of Pressure (COP) – time<br>- Centre of Pressure (COP) – position<br>(all during sit-to-walk task)<br><u>Measuring moments:</u><br>Pre- and post-intervention, 4-week and 12-week follow-up<br><u>Between-group differences:</u><br>Between-group differences were found in favour of the action observation group at all three measurement moments for NFOGQ, UPDRS III, PDQ39 – mobility, and PDQ39 – bodily discomfort. Between-group differences were also found in favour of the action observation group for PDQ39 – total and UPDRS II (4 and 12 weeks), BBS (4 weeks), and the 6-min walking test (12 weeks). Significant differences were found in the COP profiles. |
|  | <u>Action observation plus sonification:</u><br>n = 12; 74.7 yrs (SD: 5.9) | Action observation plus sonification was applied. Participants watched eight videos with eight different gait-related movements (e.g. weight shifting). Each video was composed of images and sounds (obtained with sonification). Afterwards, patients performed the actions for the same time twice (once without watching and once while watching again). | 16 sessions of 60 min in 8 weeks |  |
|  | <u>Cue control:</u><br>n = 10; 72 yrs (SD: 5.8) | The same eight motor gestures were performed also in the cue control group with the same order and amount of time by using attentional strategies (e.g. stripes on the floor on beat of the metronome). | 16 sessions of 60 min in 8 weeks |  |

| Reference | Population | Intervention and task | Amount of supervised practice | Measurement instruments, moments, and outcome |
| --- | --- | --- | --- | --- |
| Pelosin et al., 2010 | N = 18 persons with PD | <u>Task:</u> Strategies useful in circumventing freezing of gait. |  | <u>Measurement instruments:</u><br>- Freezing of Gait Questionnaire (FOG-Q)<br>- Freezing of Gait (FOG) diary (number of FOG episodes, three conditions: start, turn, and obstacle)<br>- Timed Up and Go Test<br>- 10-Metre Walking Test<br>- Tinetti Scale<br>- 39-item PD questionnaire<br><u>Measuring moments:</u><br>Pre- and post-intervention, 4-week follow-up<br><u>Between-group differences:</u><br>Between-group differences were found in favour of the action observation group for the FOG-Q (at follow-up) and the number of FOG episodes at follow-up (overall and for the start condition). |
|  | <u>Action observation:</u><br>n = 9; 68.8 (SD: 4.1) | Participants watched six 6-min video clips twice each showing strategies to prevent FOG. After observation, patients were asked to practice (for the remaining time of the session – 36 minutes). | 12 sessions of 60 min in 4 weeks |  |
|  | <u>Landscape observation:</u><br>n = 9; 70.2 (SD: 6.8) | Participants watched videos with landscape pictures for the same amount of time. They performed the same movement as the experimental group. | 12 sessions of 60 min in 4 weeks |  |
| Pelosin et al., 2018 | N = 64 persons with PD | <u>Task:</u> Strategies useful in circumventing freezing of gait (FOG). |  | <u>Measurement instruments:</u><br>- A new FOG questionnaire<br>- Timed Up and Go Test<br>- 10-Metre Walking Test<br>- Berg Balance Scale<br><u>Measuring moments:</u><br>Pre- and post-intervention, 4-weeks follow-up<br><u>Between-group differences:</u><br>Between-group differences were not reported. |
|  | <u>Action observation:</u><br>n = 32; 70.4 (SD: 4.5) | Participants watched six 6-min video clips twice each showing strategies to prevent FOG. After observation, patients were asked to practice (for the remaining time of the session – 36 minutes). | 10 sessions of 45 min in 5 weeks |  |
|  | <u>Landscape observation:</u><br>n = 32; 72.8 (SD: 3.1) | Participants watched videos with landscape pictures for the same amount of time. They performed the same movement as the experimental group. | 10 sessions of 45 min in 5 weeks |  |

| Reference | Population | Intervention and task | Amount of supervised practice | Measurement instruments, moments, and outcome |
| --- | --- | --- | --- | --- |
| Franceschini et al., 2012 | N = 102 persons after stroke in the subacute phase | <u>Task</u>: 20 different daily routine tasks carried out with the upper limb (e.g. drinking from a glass, combing hair). | All participants underwent inpatient rehabilitation with at least 3 hours/day of physiotherapy. | <u>Measurement instruments</u>:<br>- Frenchay Arm Test<br>- Fugl-Meyer Test<br>- Box and Block Test<br>- Ashworth Scale – Shoulder<br>- Ashworth Scale – Elbow<br>- Ashworth Scale – Wrist<br>- Functional Independence Measure – Motor Score<br><u>Measurement moments</u>:<br>Pre- and post-intervention, 4-5 months follow-up<br><u>Between-group differences</u>:<br>Differences in favour of the action observation group were found on Box and Block Test post-intervention and at follow-up. |
|  | <u>Effect of action observation</u>:<br>n = 53; 67.0 yrs (SD: 12.4) | Participants watched a video of one of the 20 tasks and imitated the action under supervision and with verbal feedback and assistance if needed. | 20 sessions of 2 x 15 min in 4 weeks |  |
|  | <u>'Sham' observation</u>:<br>n = 49; 65.7 yrs (SD: 11.9) | Participants were shown 5 static images displaying objects and executed a cognitive task. They performed limb movements according to a standard sequence, simulating those performed by the EG, involving shoulder and elbow joint mobilisation. | 20 sessions of 2 x 15 min in 4 weeks |  |
| Mancuso et al., 2021 | N = 36 persons with stroke in acute phase | <u>Task</u>: Functional activities with the upper limbs.<br>All participants received conventional therapy with a range of different patient-tailored interventions, not involving AOT or TOT objects. | All subjects underwent rehabilitative treatment for 4 consecutive weeks, 5 days per week. Each session consisted of 60 min. | <u>Measurement instruments</u>:<br>- Fugl-Meyer Assessment for Upper Extremity<br>- Box and Block Test<br>- Functional Independence Measure (FIM)<br>- Modified Ashworth Scale<br><u>Measurement moments</u>:<br>Pre- and post-intervention<br><u>Between-group differences</u>:<br>Differences in favour of the action observation group were found for the FIM. |
|  | <u>Action observation training (AOT)</u>:<br>n = 16; median: 64.5 yrs (IQR: 15.8) (mean/SD: NR) | Participants observed 20 different videos of daily activities carried out with the upper limb and performed the activities afterwards (3 min of observation and 2 min of action performance for 3 motor sequences, repeated twice). | 20 sessions of 30 min for 4 wks (additional to the rehabilitative treatment) |  |
|  | <u>Task-oriented training (TOT)</u>:<br>n = 16; median: 76.5 yrs (IQR: 13.7) (mean/SD: NR) | Participants performed the same functional activities with the upper limbs, in both unimanual and bimanual modalities, without watching the video beforehand. | 20 sessions of 30 min for 4 wks (additional to the rehabilitative treatment) |  |

| Reference | Population | Intervention and task | Amount of supervised practice | Measurement instruments, moments, and outcome |
| --- | --- | --- | --- | --- |
| Sale et al., 2014 | N = 67 persons after stroke in the subacute phase<br>66.5 yrs (SD: 12.7) | <u>Task</u>: Functional upper limb activities. | All subjects underwent in-patient rehabilitation consisting of at least 3 hours/day | <u>Measurement instruments</u>:<br>- Fugl-Meyer Test (FMT) – all subjects<br>- Fugl-Meyer Test – right hemiparetic subjects<br>- Fugl-Meyer Test – left hemiparetic subjects<br>- Box and Block Test (BBT)-all subjects<br>- Box and Block Test – right hemiparetic subjects<br>- Box and Block Test – left hemiparetic subjects<br><u>Measurement moments</u>:<br>Pre- and post-intervention, and 4-5 months follow-up<br><u>Between-group differences</u>:<br>Between-group differences were found in favour of the action observation group for FM and BBT (post-intervention and follow-up, all subjects and left hemiparetic subjects). |
|  | <u>Action observation training</u>:<br>n = 33; data on age NR | Participants watched one of 20 videos with different routine actions carried out with the upper limb and performed the same movement afterwards. | 20 sessions of 2x15 mins for 4 weeks |  |
|  | <u>Control group</u>:<br>n = 34; data on age NR | Participants watched static images of objects and performed a cognitive task to keep their attention high. Afterwards, they performed the same movements as in the EC. | 20 sessions of 2x15 mins for 4 weeks |  |
| Rojasav et al., 2020 | N = 33 elderly persons with mild cognitive impairment | <u>Task</u>: Walking. |  | <u>Measurement instruments</u>:<br>- Gait speed (m/s)<br>- Stride time variability (% coefficient of variation)<br>- Stride length variability (% coefficient of variation)<br>in both single- and dual-task conditions<br><u>Measurement moments</u>:<br>Pre- and post-intervention, 4 wks follow-up<br><u>Between-group differences</u>:<br>Between-group differences between the AOGT and CT groups were found in favour of the AOGT group for gait speed during single-task and dual-task conditions post-intervention and at follow-up. |
|  | <u>Action observation with gait training (AOGT)</u>:<br>n = 11; 67.6 yrs (SD:4.6) | Participants watched a video of walking acted out by a normal healthy individual followed by gait training. | 12 sessions of 65 mins for 4-6 weeks |  |
|  | <u>Gait training (GT)</u>:<br>n = 11; 67.5 yrs (SD: 5.6) | Participants watched a video of abstract pictures followed by the same gait training as in the experimental group. | 20 sessions of 15 mins for 4-6 weeks |  |
|  | <u>Control group (CT)</u>:<br>n = 11; 65.7 yrs (SD: 2.5) | Participants received no training programme but were educated about dementia on the screening day. | 1 session |  |

N = number of participants in total, n = number of participants per group, SD = standard deviation, NA = not applicable, NR = not reported, FIM = Functional Independence Measure, AFG = absolute functional gain, FOG = Freezing Of Gait, UPDR = Unified Parkinson's Disease Rating Scale, PDQ = Parkinson's Disease Questionnaire, NFOGQ = New Freezing of Gait Questionnaire, COP = Centre of Pressure, BBS = Berg Balance Scale, FOG-Q = Freezing of Gait Questionnaire, FIM = Functional Independence Measure, FMT = Fugl-Meyer Test, BBT = Box and Block Test, AOGT = Action observation with gait training group, CT = control group

### Dual-task learning

Forty-three studies were included, with a sample size ranging from 12 to 134. Three included persons after stroke, of which two studies^62,63^ practised walking and one study^64^ practised daily life activities. Three studies included orthopaedic patients, who all practised balance exercises.^65–68^ Five studies included persons with dementia, and all practised walking and/or balance tasks.^69–73^ Six studies included people with Parkinson’s, of which two studies focused on balance training,^74,75^ three practised walking,^76–78^ and one practised aquatic exercises.^21^ Seventeen studies included older adults, of which 13 practised walking and/or balance exercises^79–91^ and the other studies practised aerobic exercises,^92^ stepping exercises,^93^ resistance training,^94^ stationary biking,^95^ or agility and strength training.^96^ Three studies included older adults with balance impairments and practised walking and/or balance.^24,96–98^ One study included older adults with fall histories and practised walking and balance.^99^ Three studies included older adults with cognitive impairments and practised walking and/or balance^100,101^ or dancing.^102^

Within the dual-task conditions, the secondary task was either a motor or a cognitive task. Secondary motor tasks included (avoiding) obstacles, carrying or playing with obstacles (e.g. a grocery bag, a tray, rattle, umbrella, or musical instruments), exercises with a ball (e.g. bouncing, passing, throwing, catching, holding, kicking) practicing daily life activities such as (un)buttoning a shirt, putting beans in a container (non-dominant hand) and unscrewing a nut and bolt, drawing a letter on the floor with one of their feet. Secondary cognitive tasks included engaging in conversations; singing; arithmetic tasks (e.g. 2-forward and 3-backward calculations); repeating animals’ names; reading words or sentences backwards; counting/reciting the days of week; simple word games (e.g. coming up with a word that starts with the last letter of the previous word or naming as many words starting with the letter P (or another random letter); remembering cards; repeating phrases; playing phonemic word chain games; reciting a poem; answering questions about the participants’ orientation to a person (identifying their name), time (date, month, or year), and place (current location); reacting to virtual situations (e.g. you’re in a taxi but do not have your wallet); explaining the order of wearing clothes (e.g. dress, skirt, shirt, tie); talking about daily routines; making a shopping list; categorisation (e.g. types of land animals, drinks, colours, objects, boys’ and girls’ names, flowers, vegetables, fruit); clock face task; alternative uses (e.g. name an object and come up with alternative uses for that object); a creativity task (e.g. name as many objects that you know that are tall); letter fluency task; planning; singing a song; comparing drawings and naming differences; word spelling (fast as possible); auditory Stroop task; remembering shapes and colours; responding to auditory cues (fast as possible); and paying attention to tripping hazards. Finally, secondary tasks presented through VR games, e.g. playing a ball game, reactive boxing game, or cleaning windows.

Eight studies observed between-group differences^70,71,77,94,101,103,104^ in favour of the dual-task learning intervention post-intervention and at the follow-up measurement (two studies with low RoB, six with some concerns about RoB, three with appropriate sample size justification). Nineteen studies^24,64–66,72–74,78,81,83,86–90,97–99,105^ observed between-group differences only immediately after the intervention (two with low RoB, 13 with some concerns about RoB, four with high RoB, 13 with appropriate sample size justifications). Eleven studies^62,63,67,69,75,79,80,84,85,93,95^ did not observe any between-group differences (all with some concerns about RoB, three with appropriate sample size justifications). Five studies^76,96,100,102,106^ did not report the between-group effects (one with low RoB, four with some concerns about RoB, three with appropriate sample size justifications).

The total study duration of these interventions ranged from 1 day^83^ up to 26 weeks.^87,92^ The total intensity of training (i.e. the number of sessions multiplied by the duration of each session) ranged from 40 minutes in one session^83^ to 4875 minutes over 65 training sessions.^92^ See Table 7 for more details.

**Table 7.**
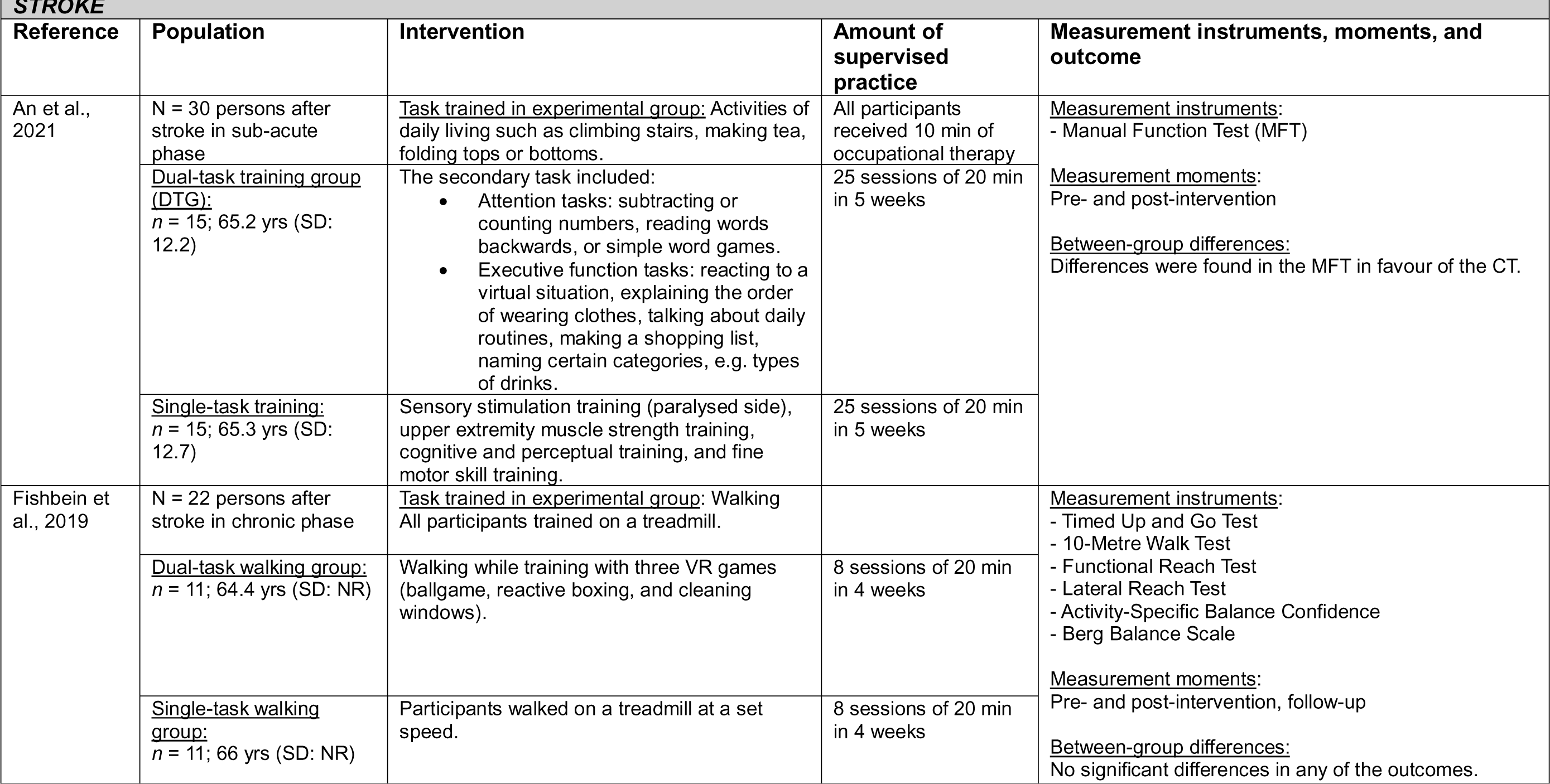
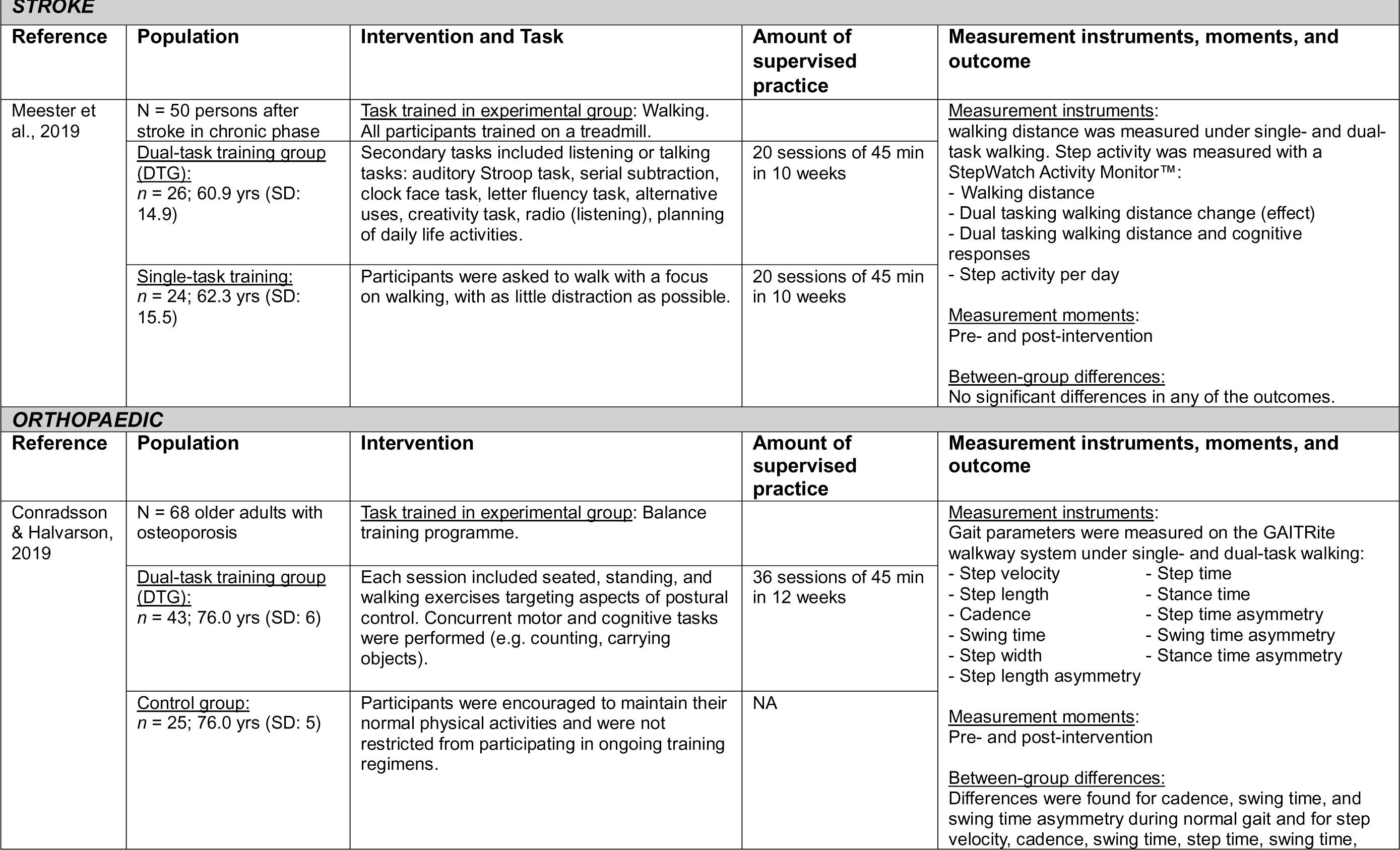
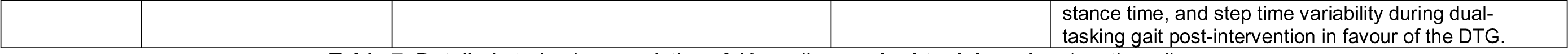
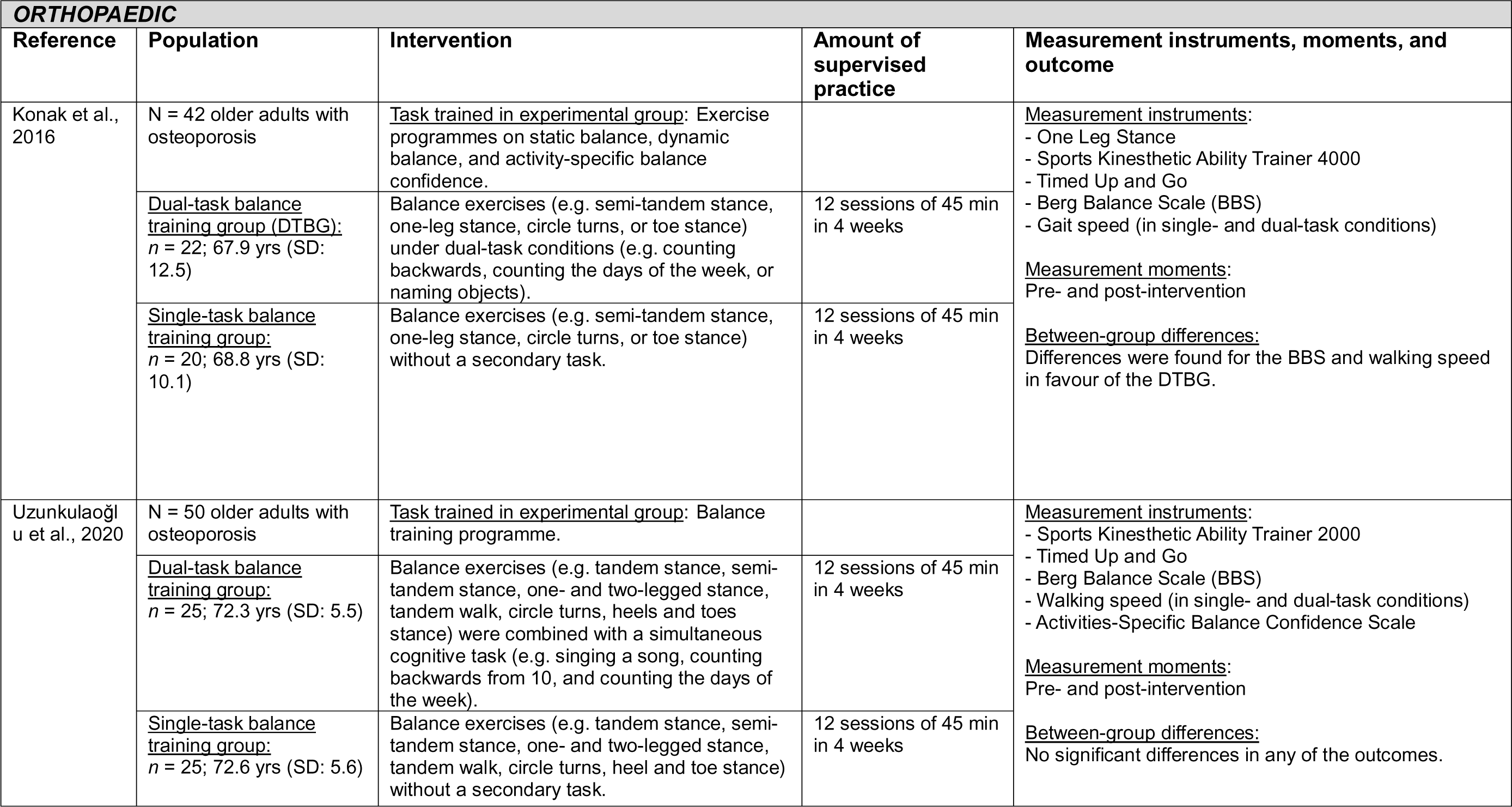
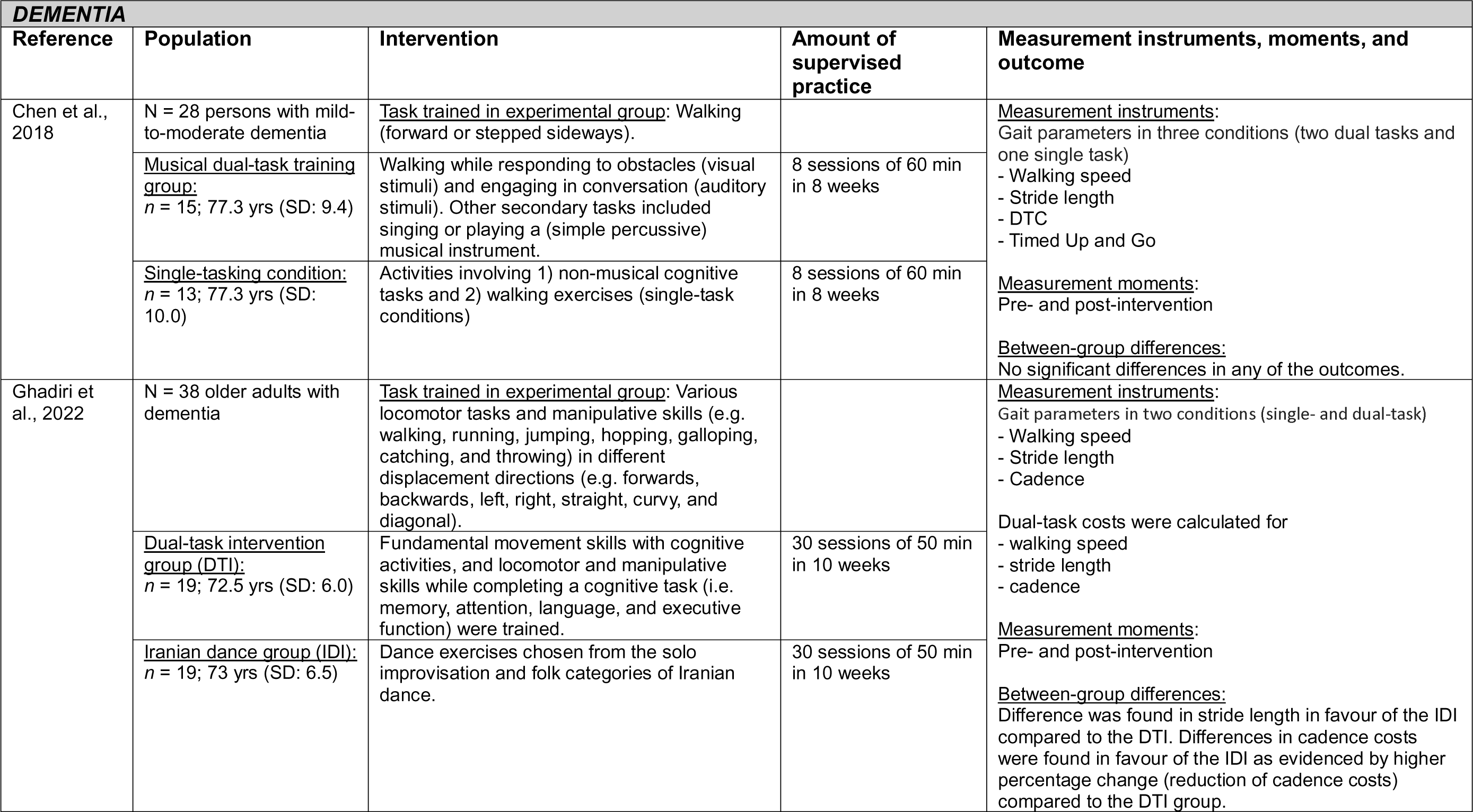
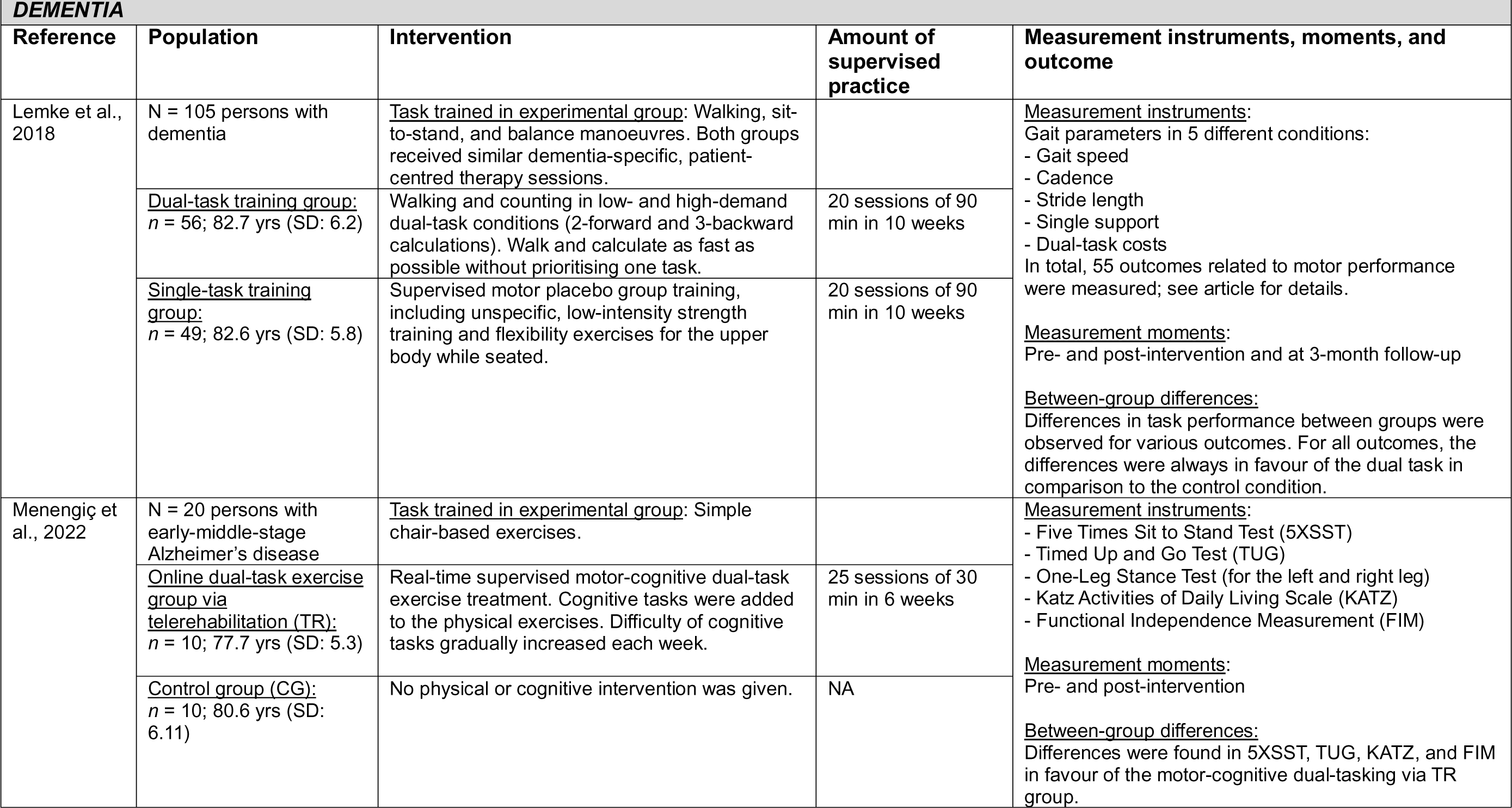
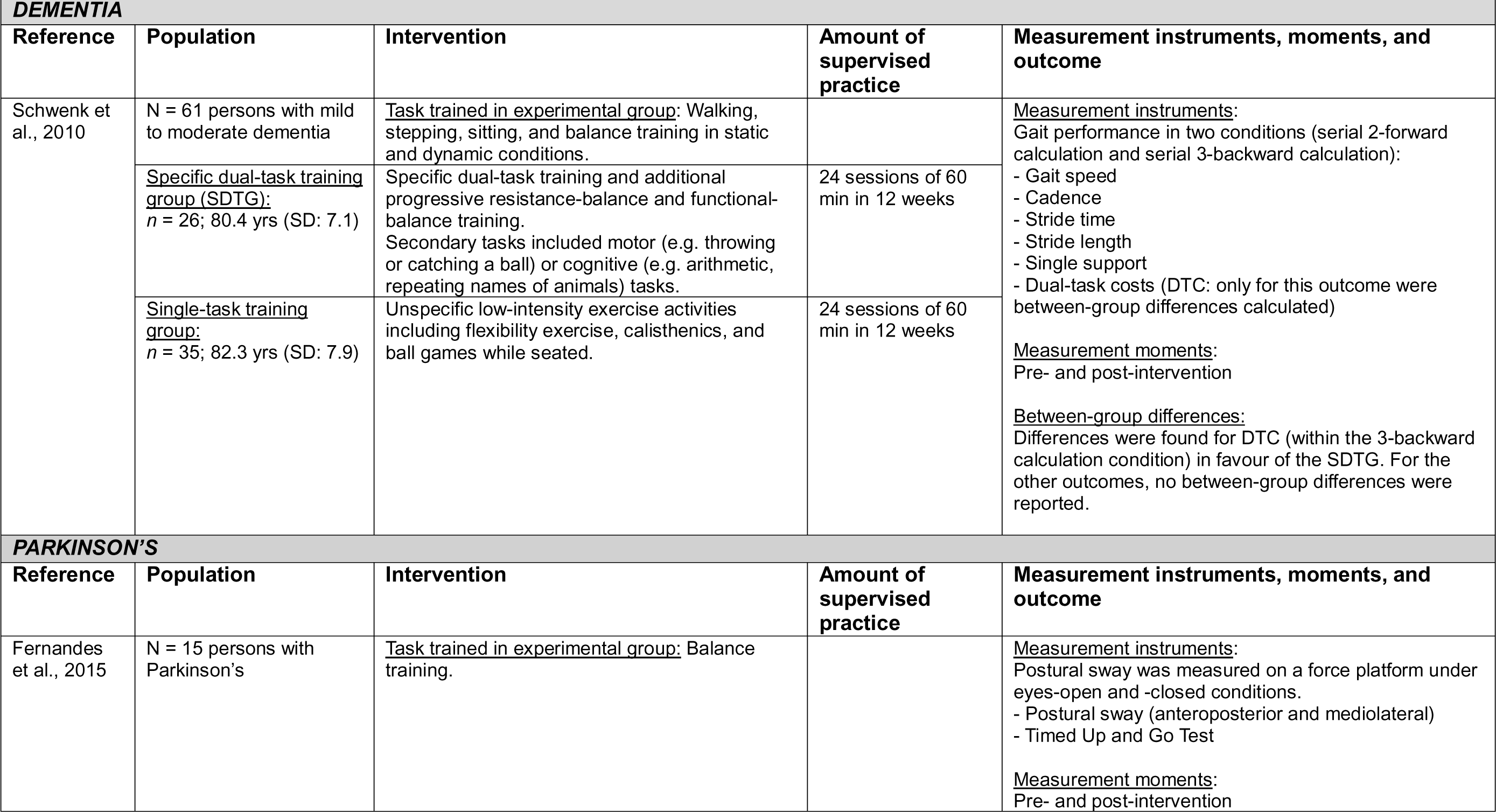
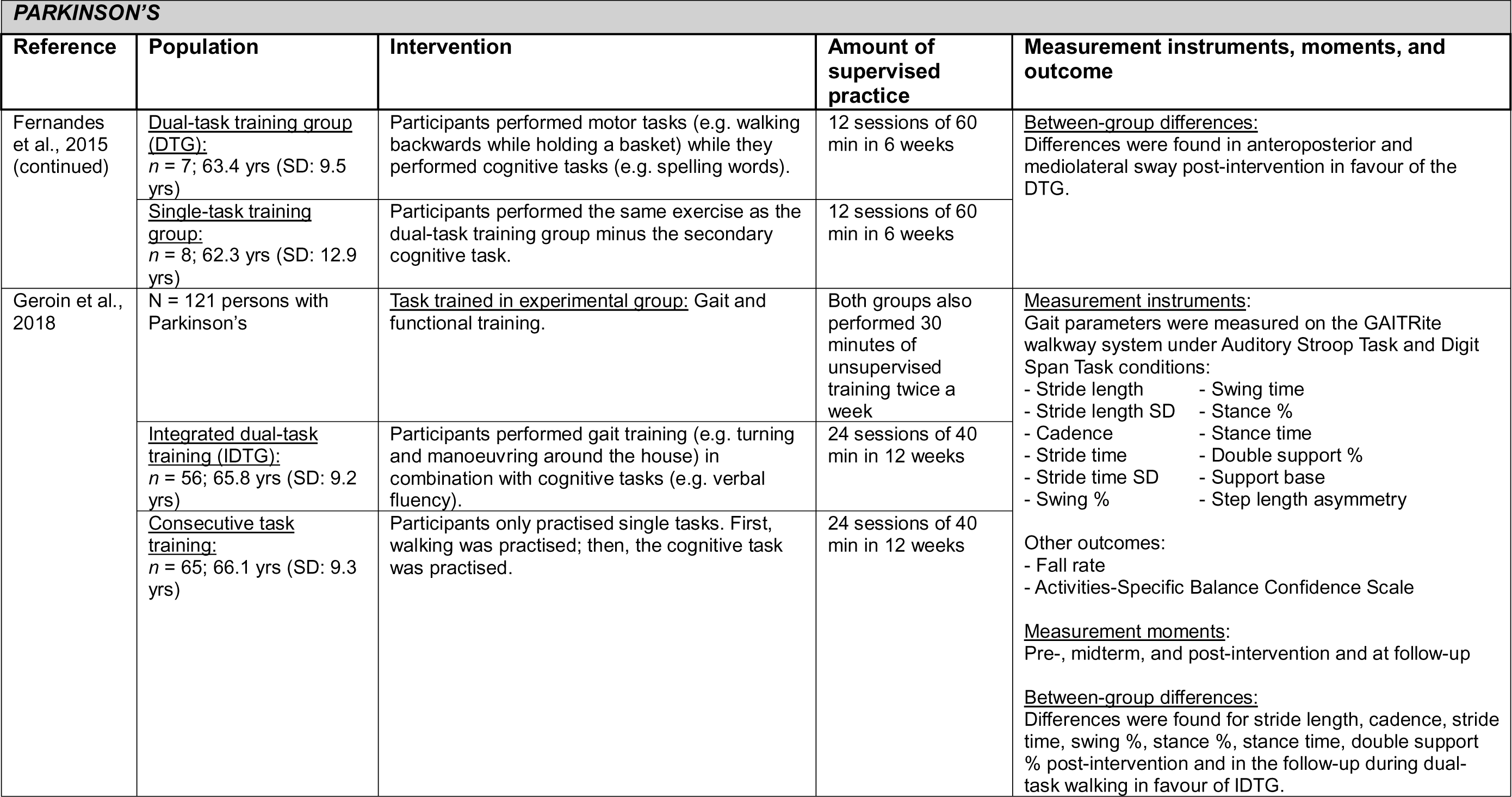
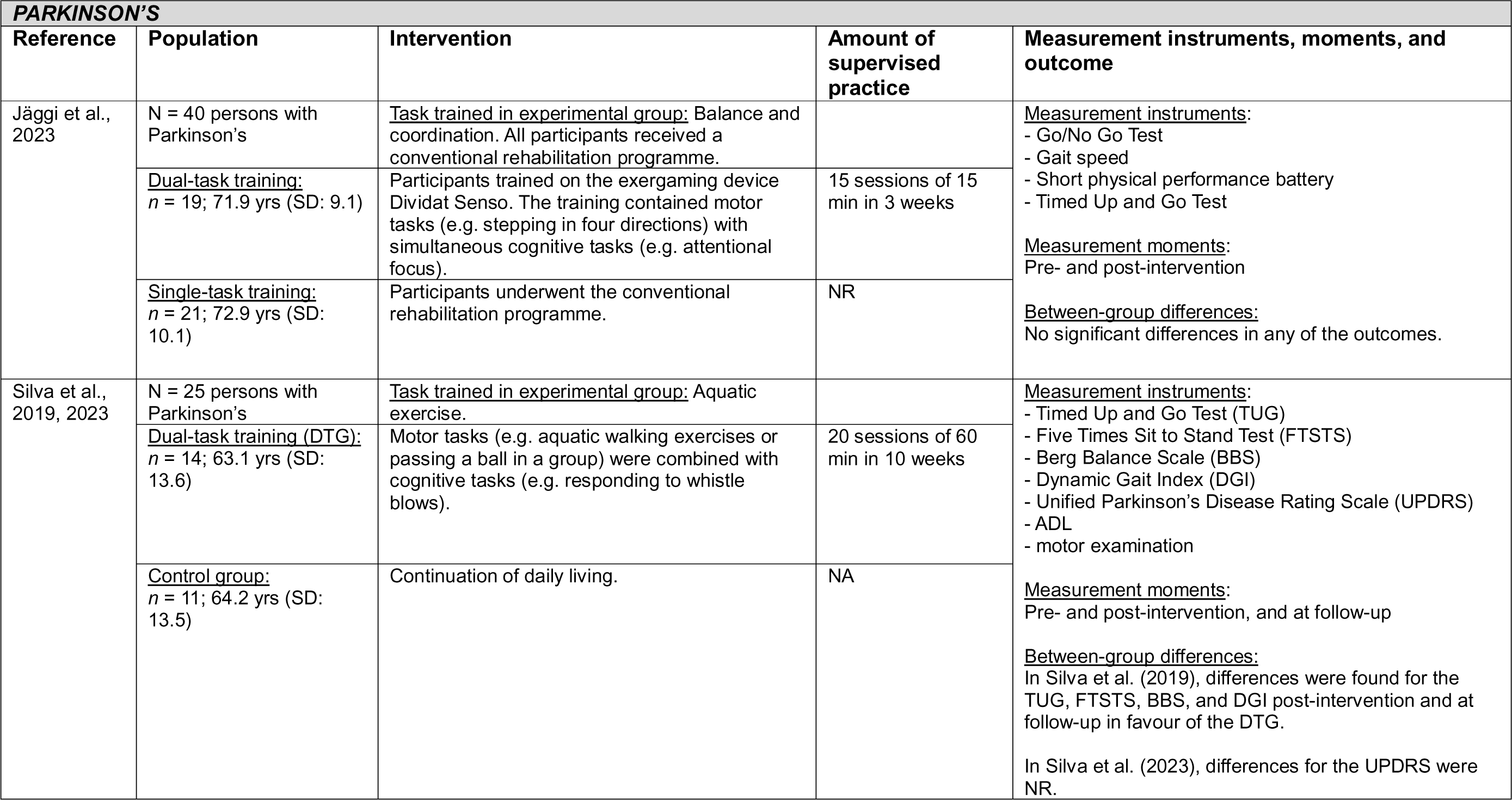
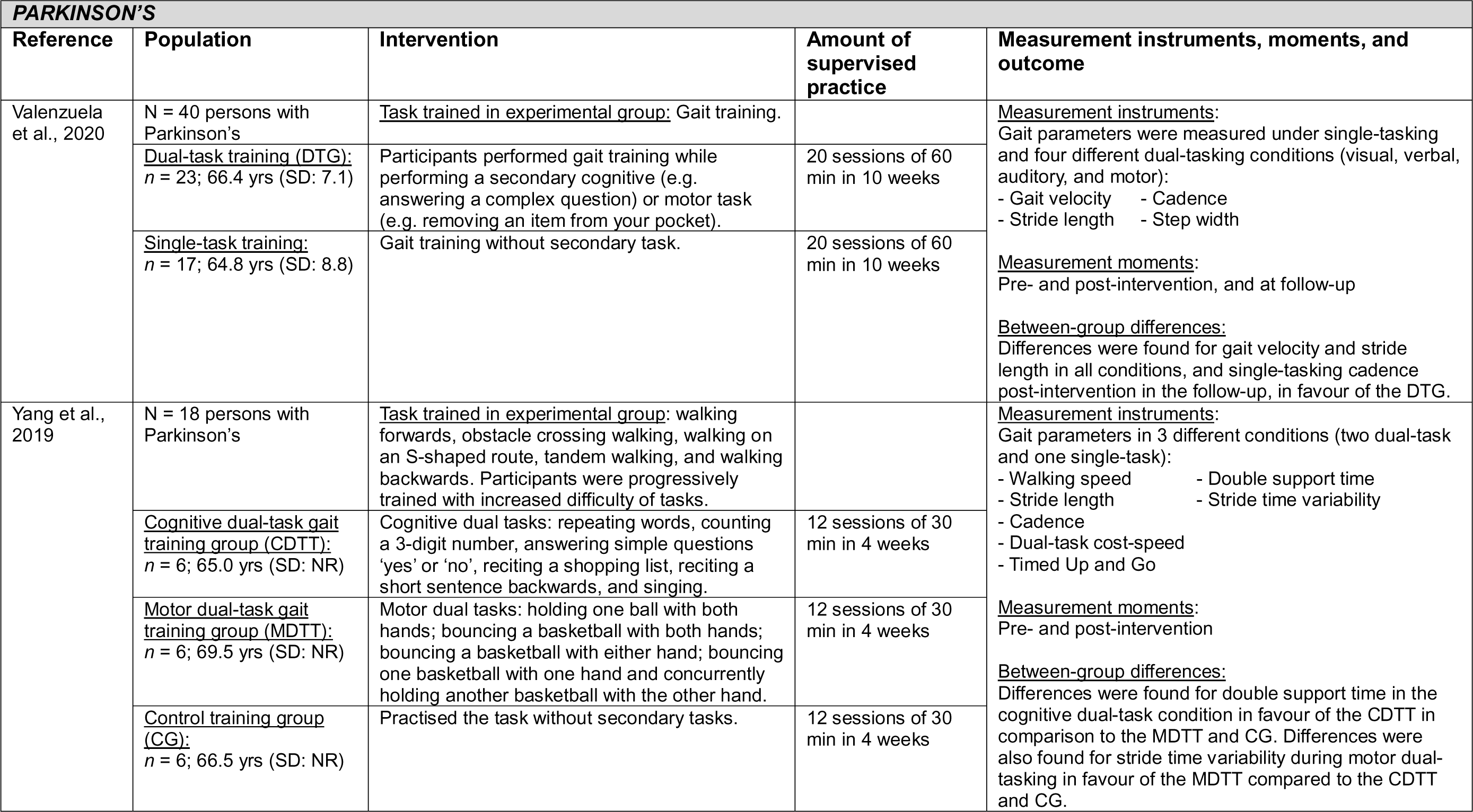
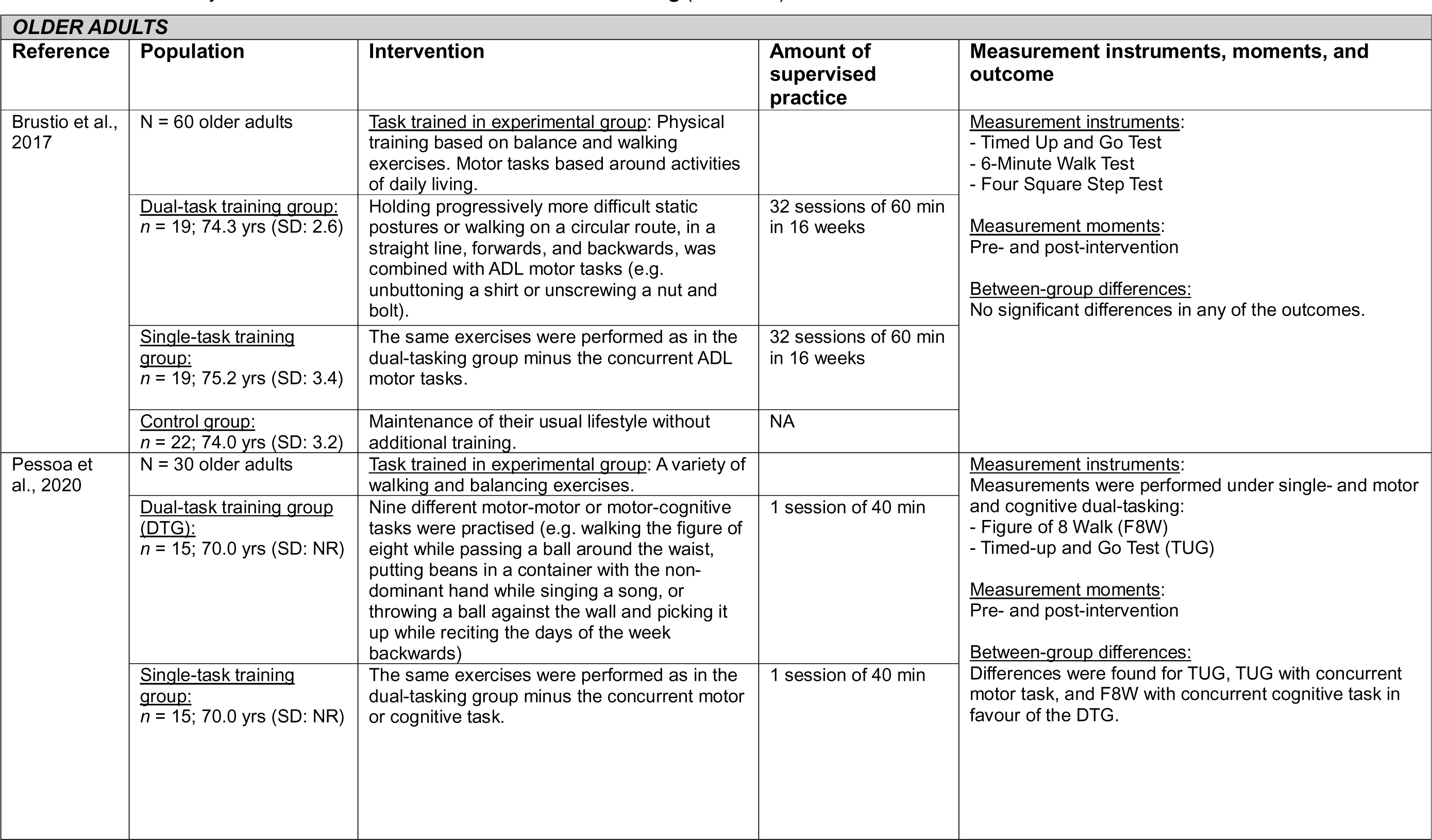
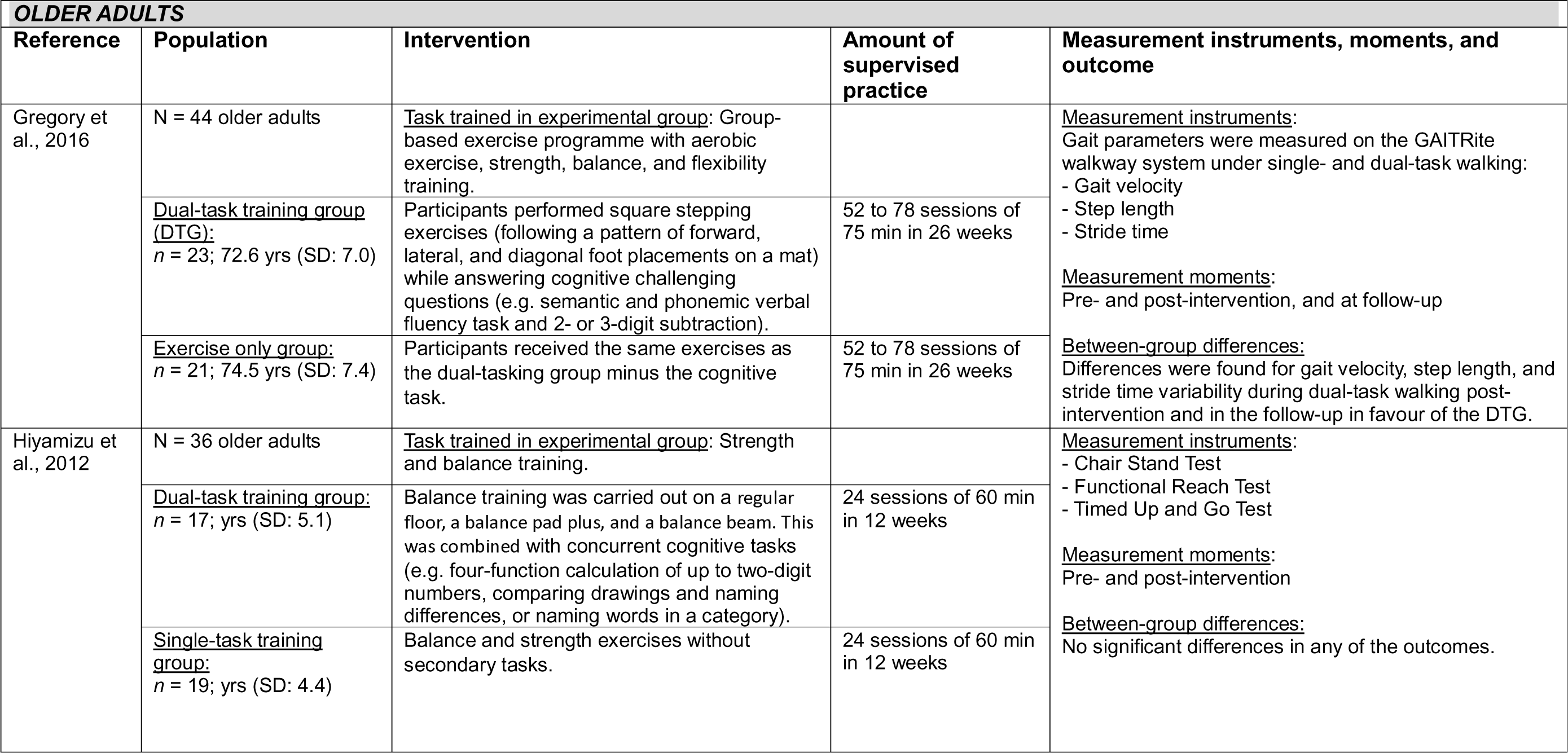
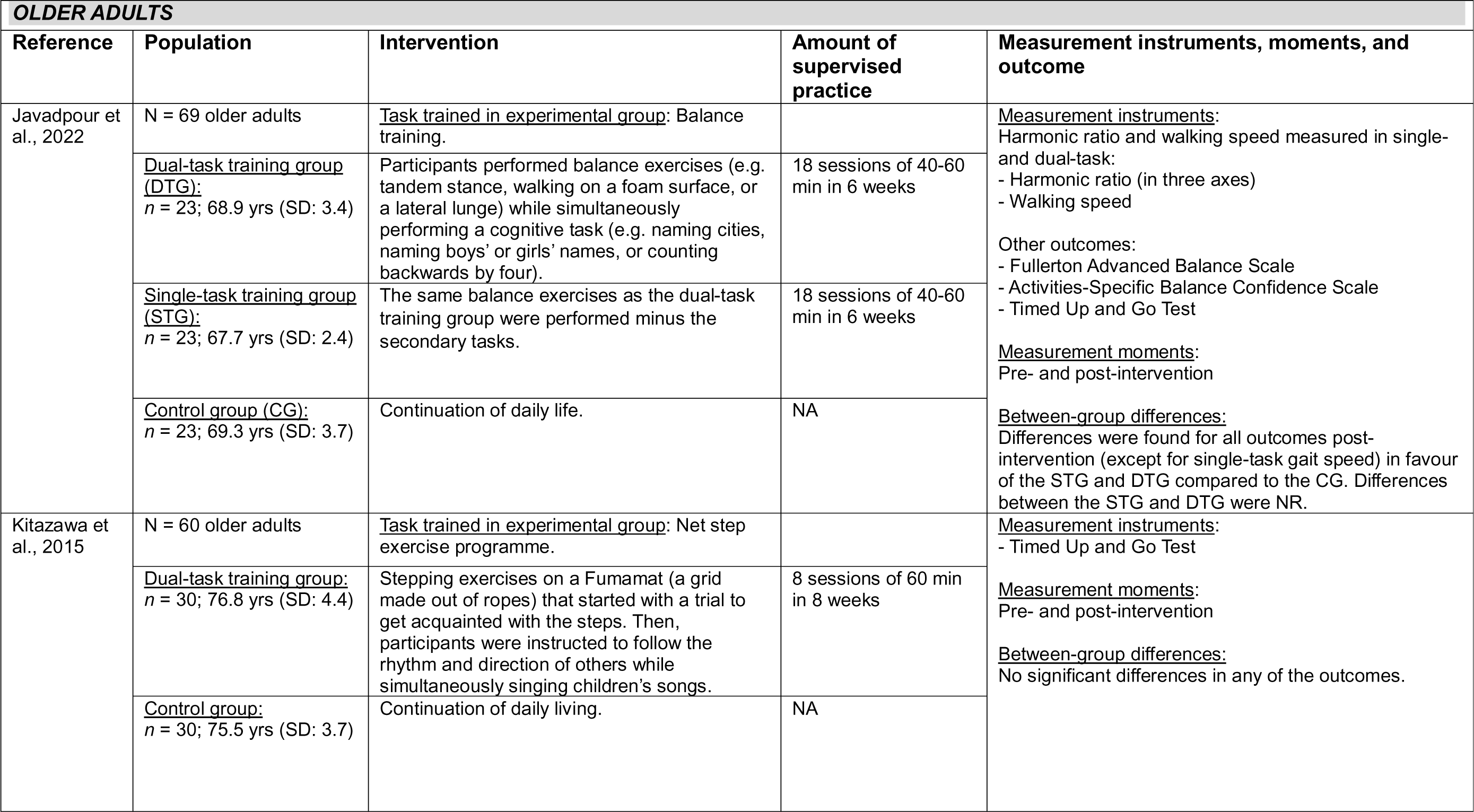
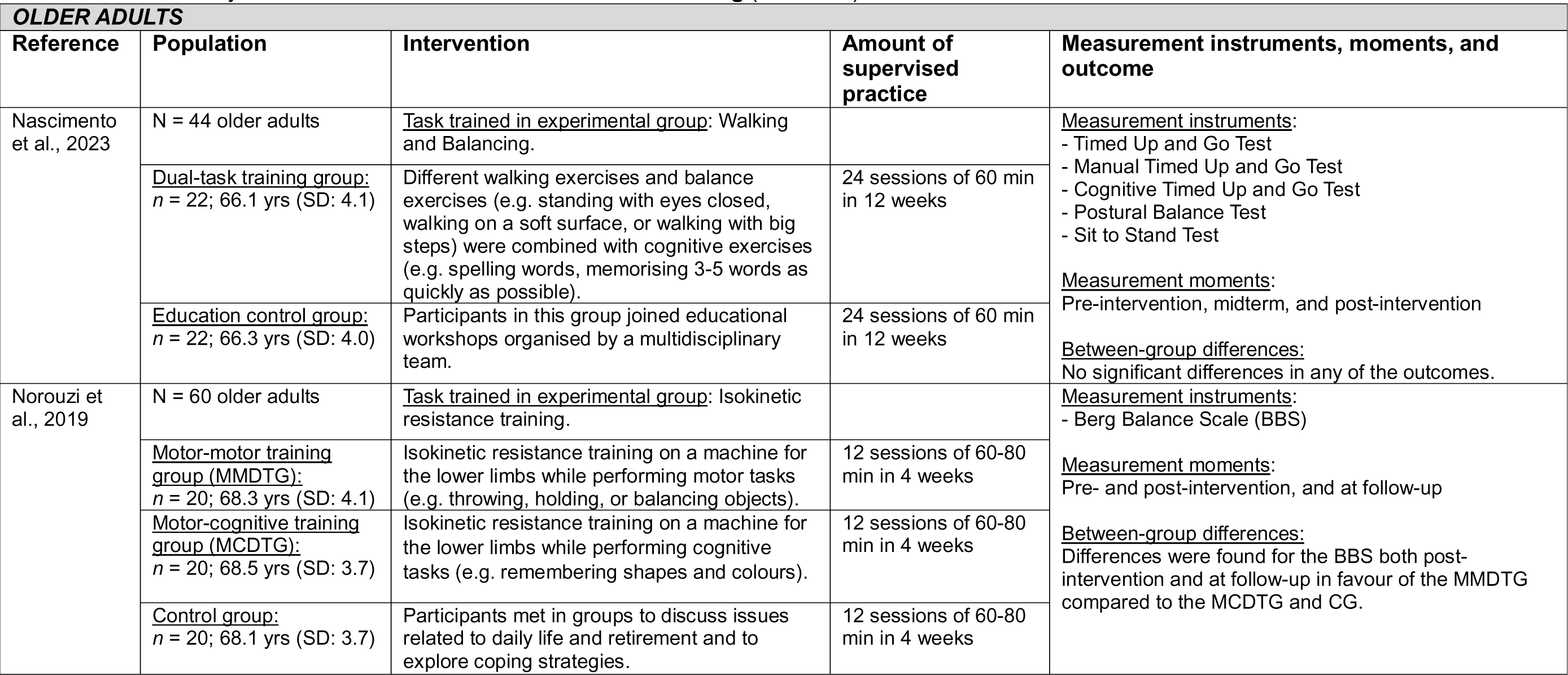
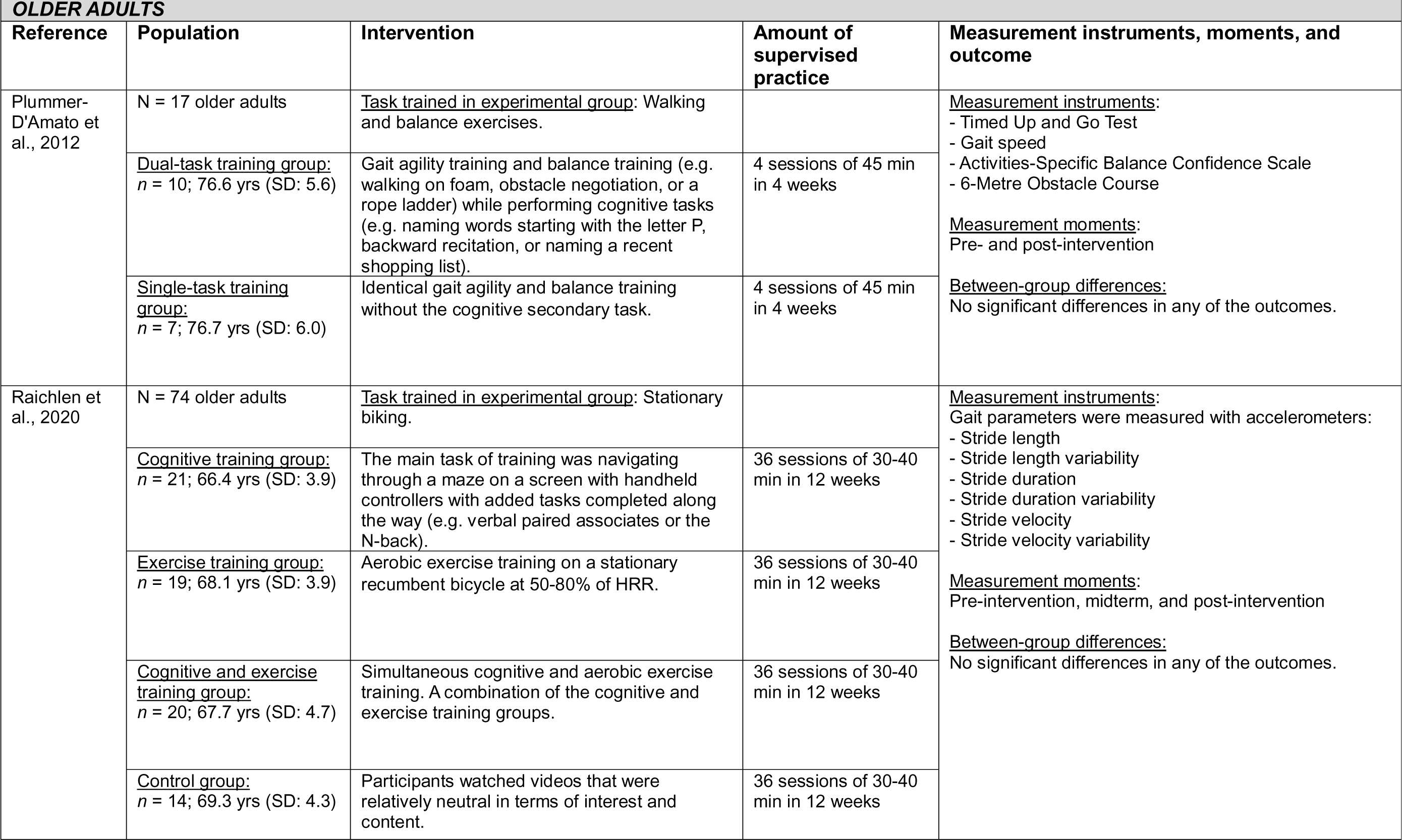
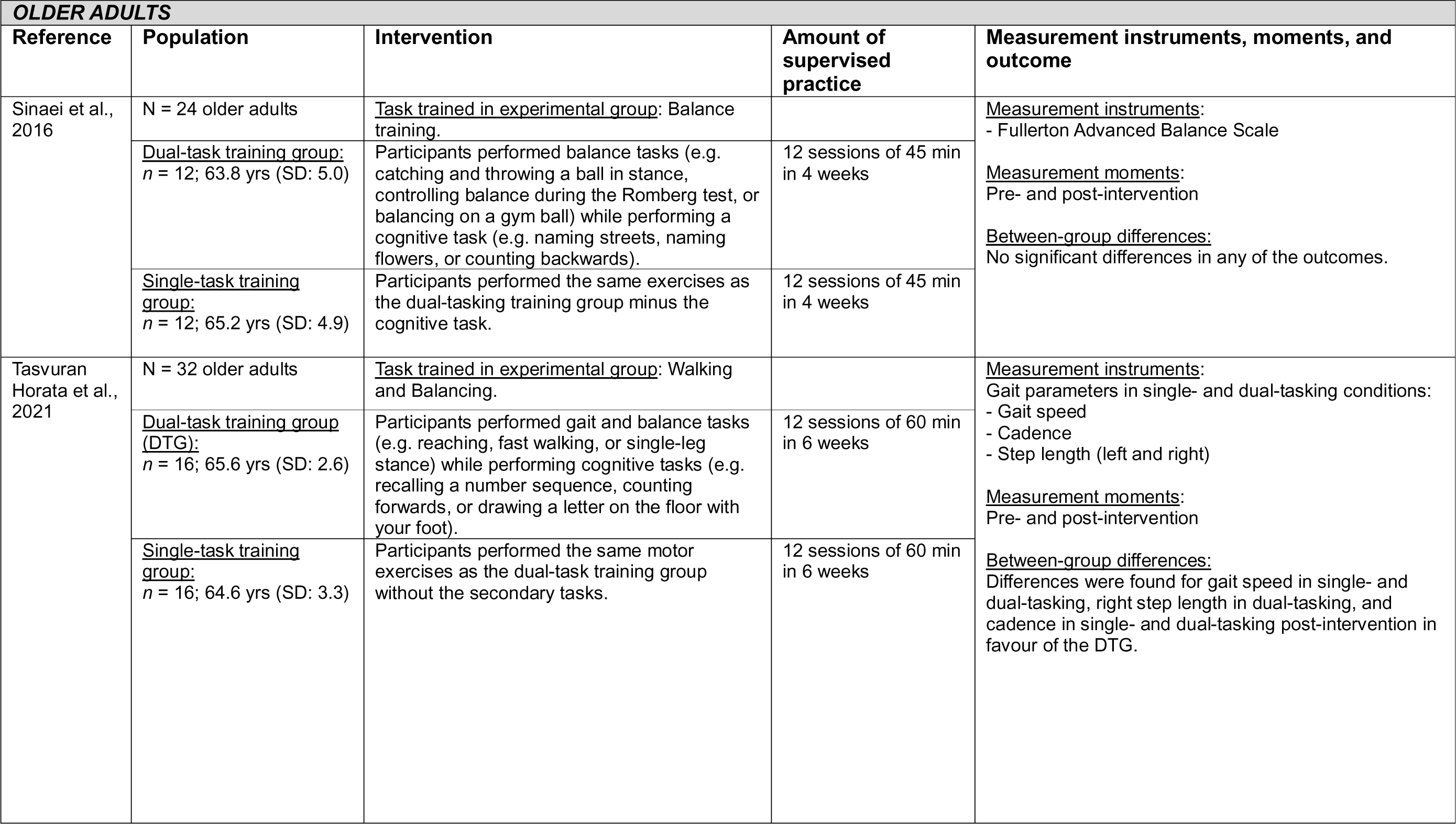
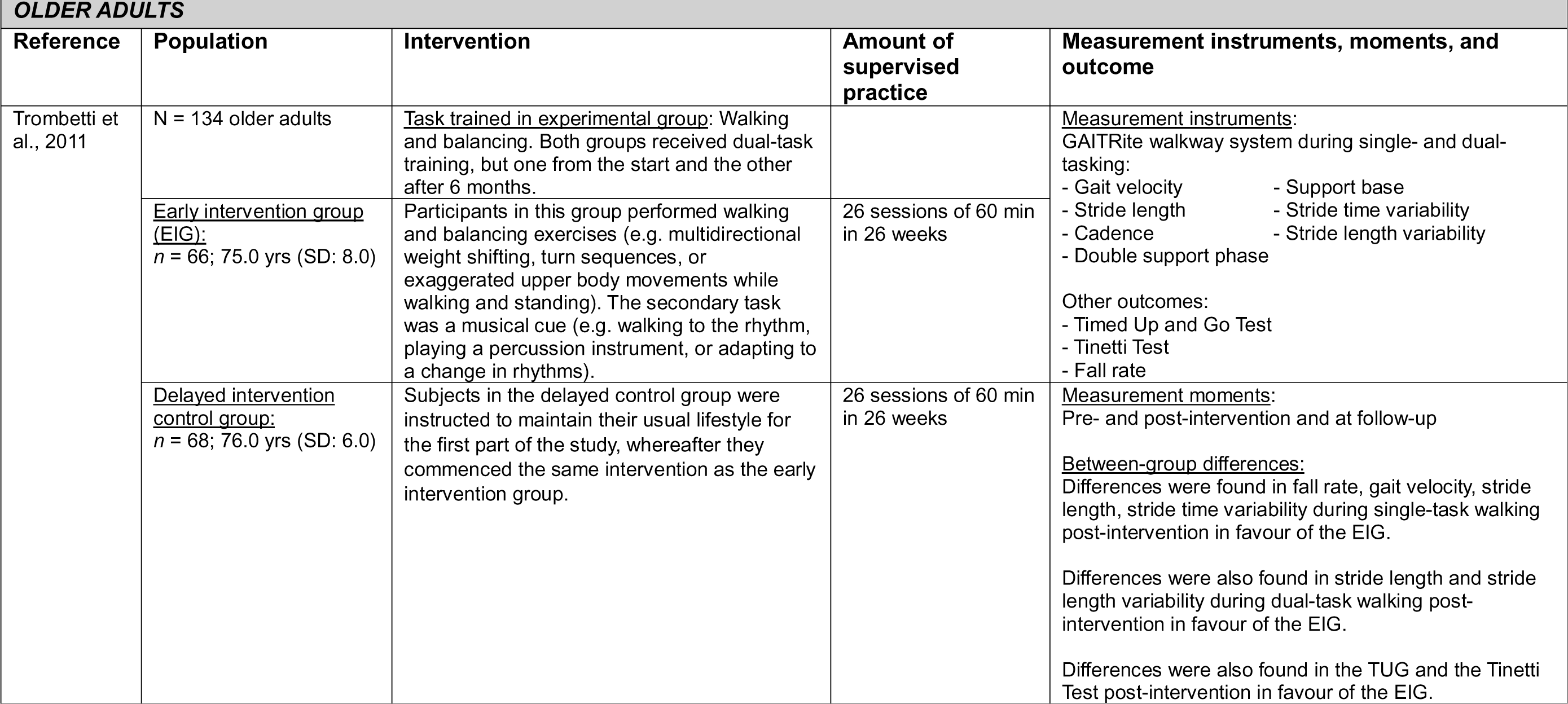
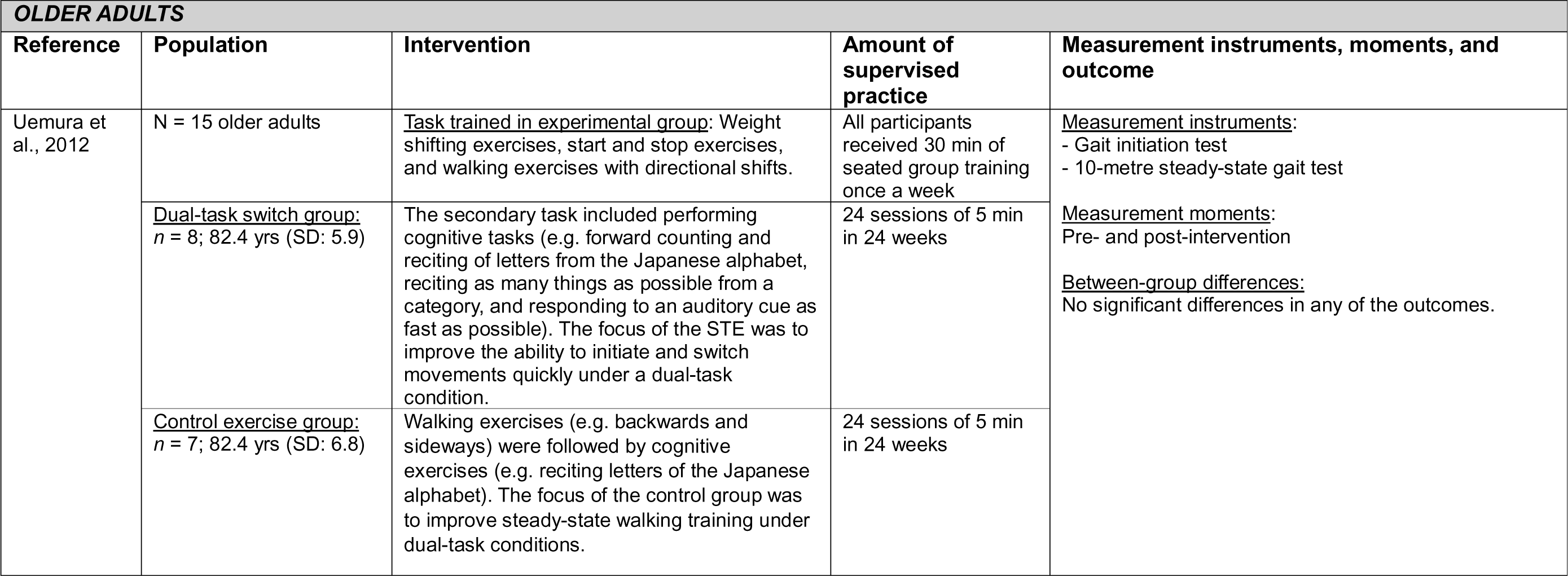
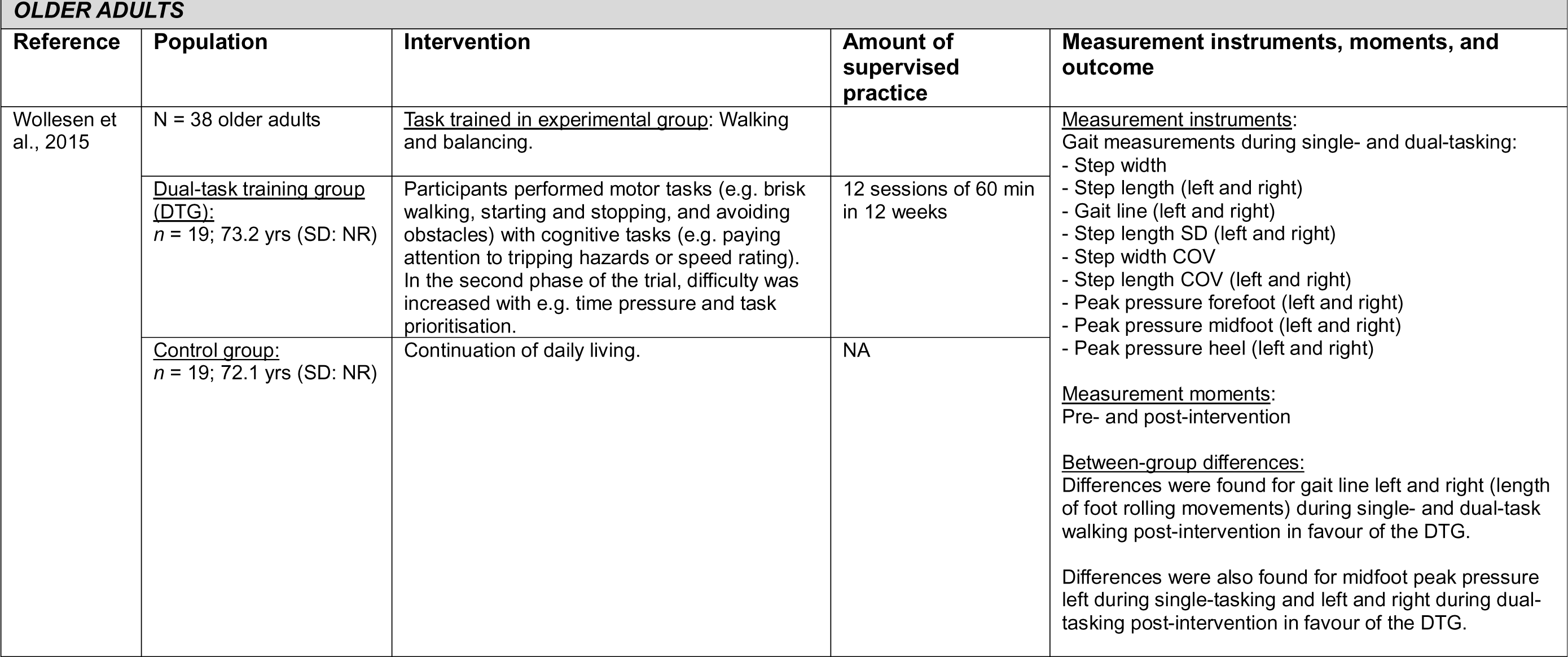
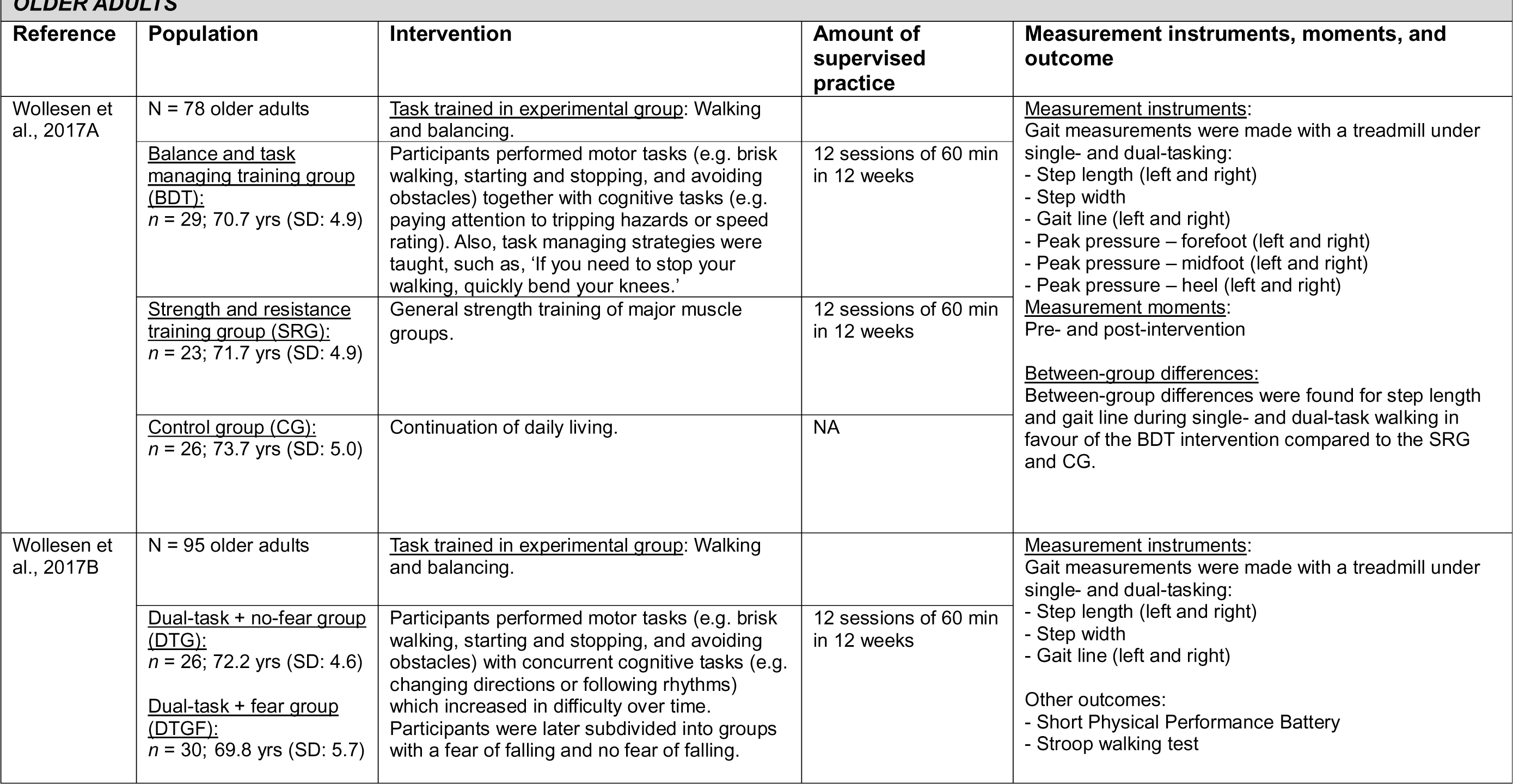
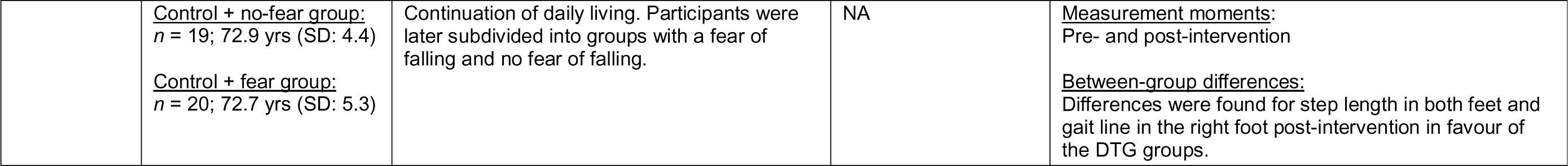
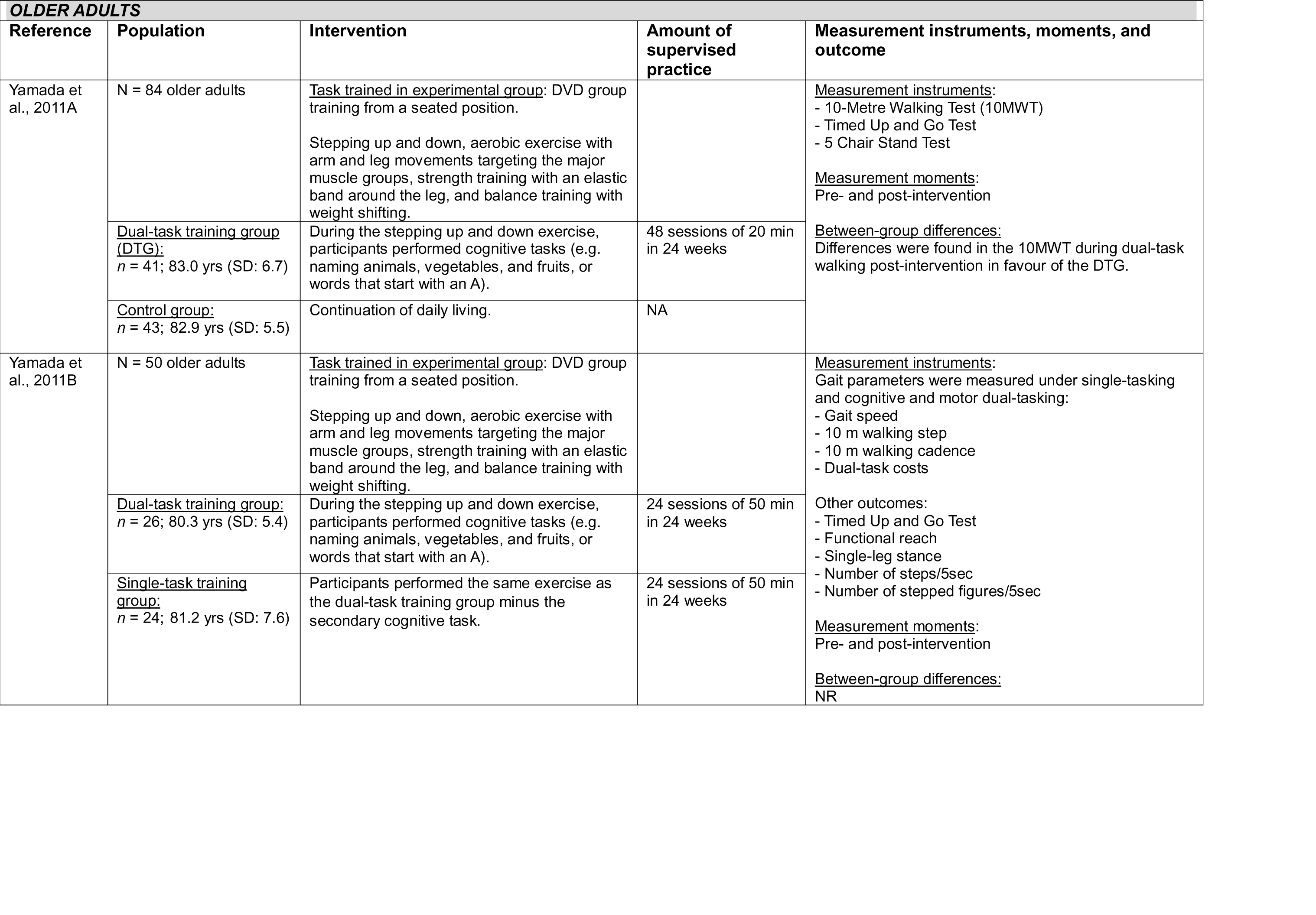
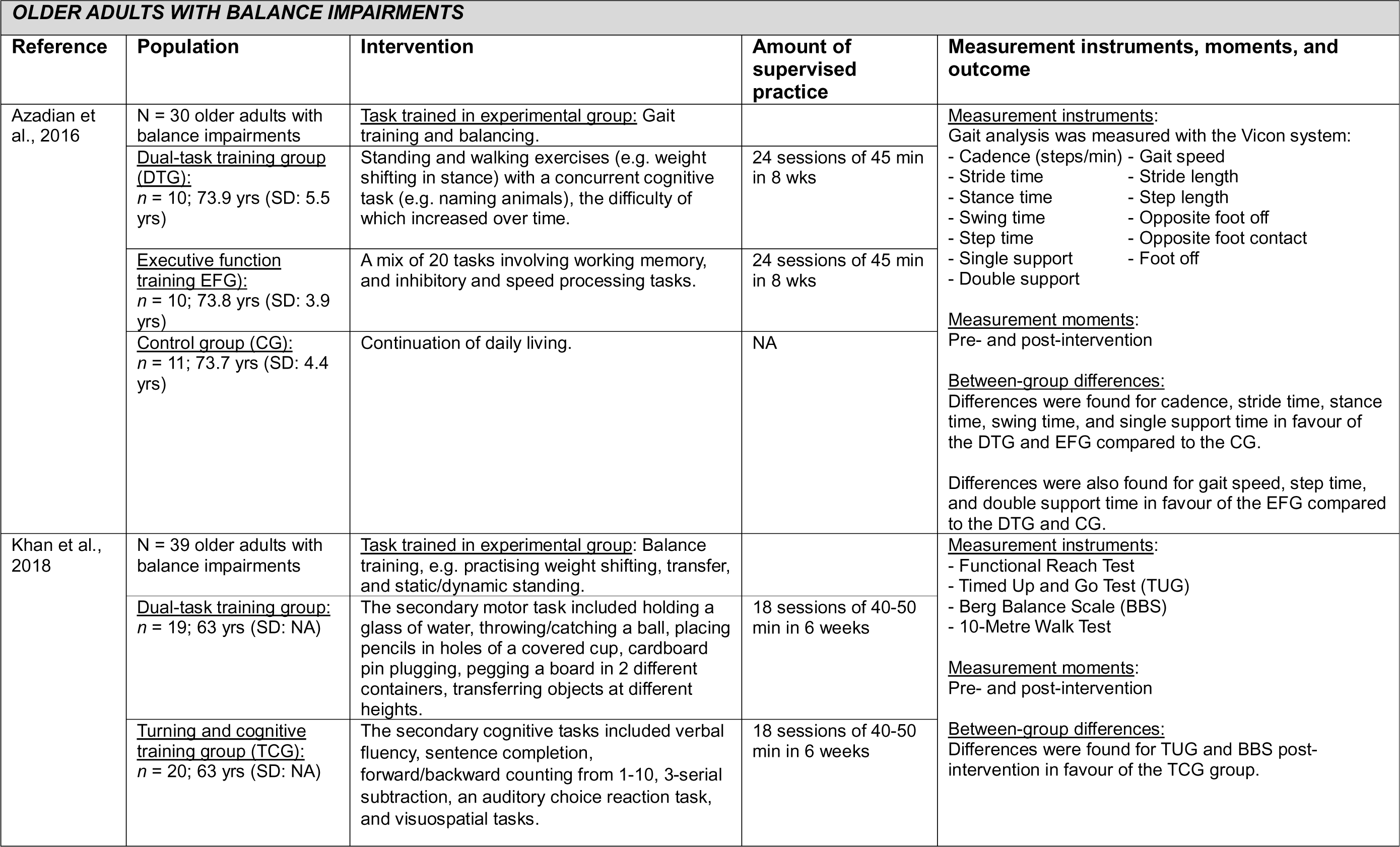
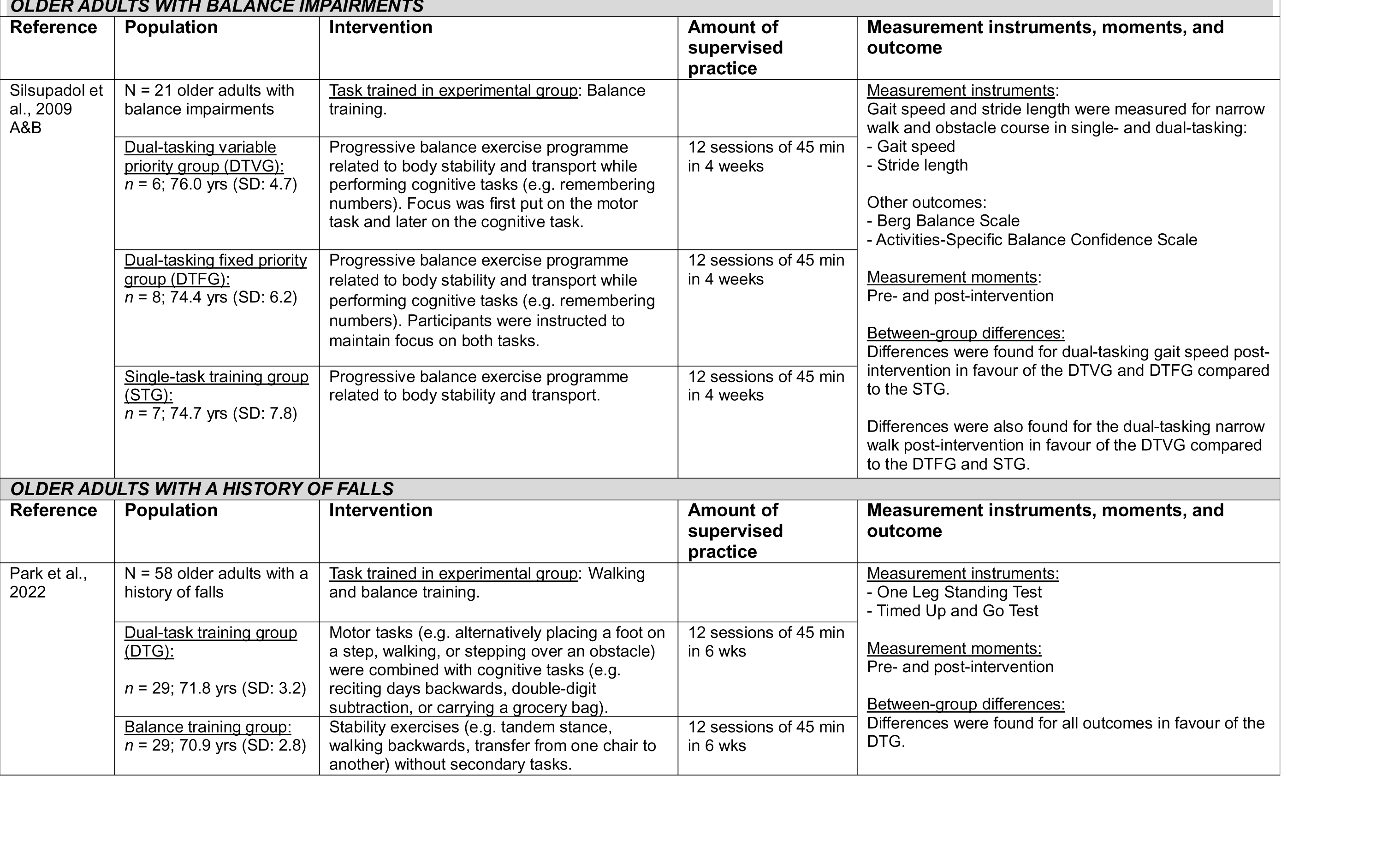
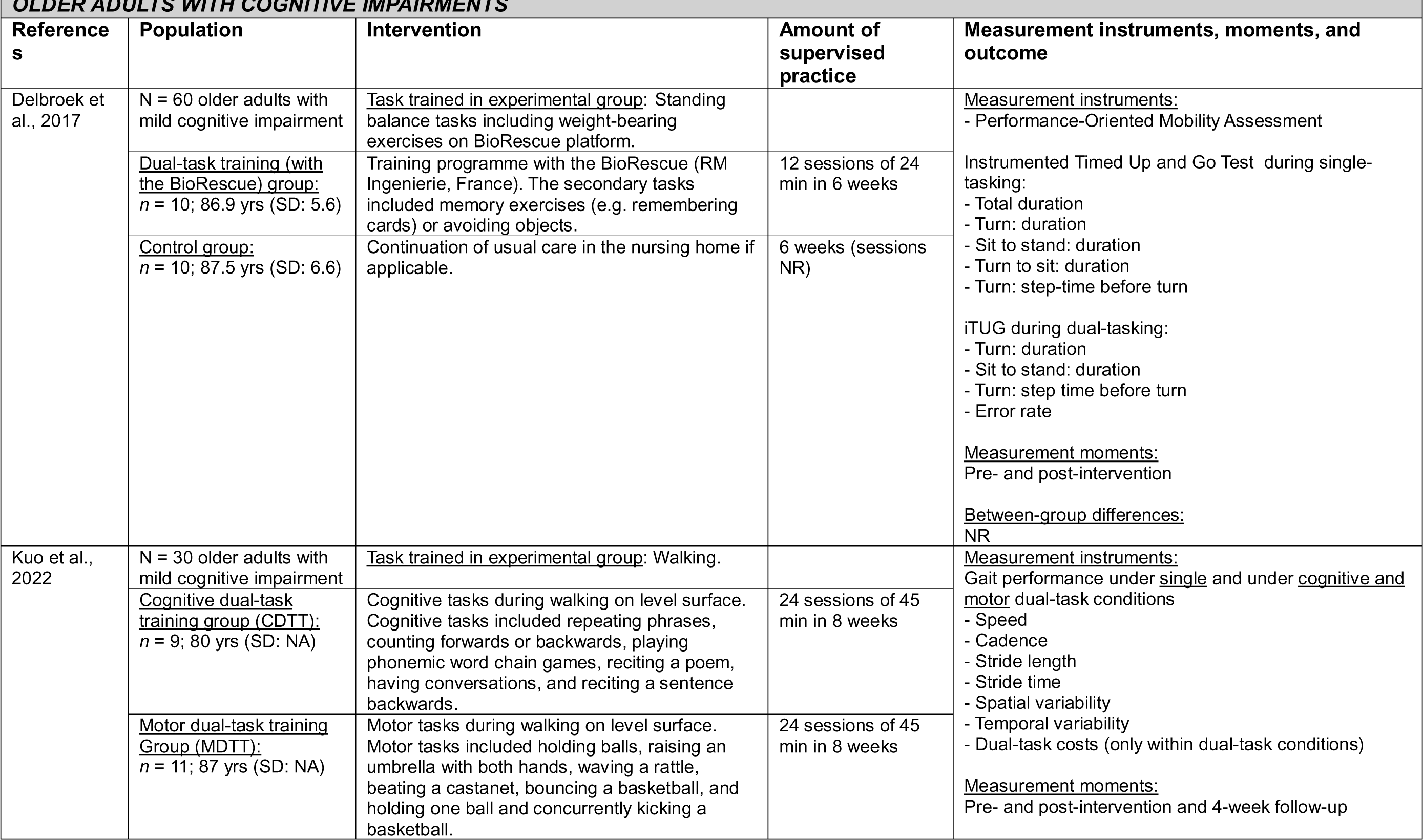
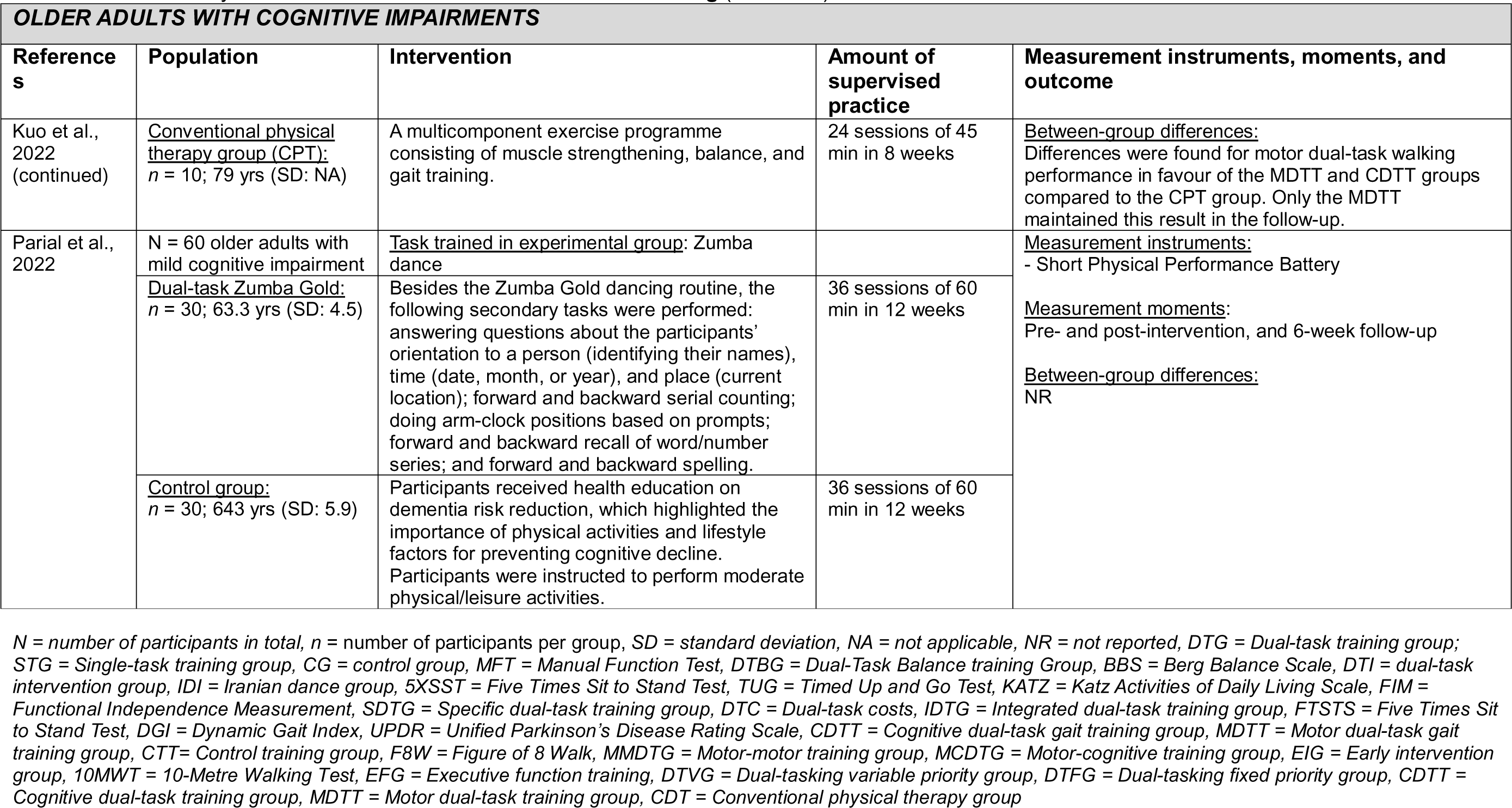
Detailed study characteristics of 43 studies on **dual-task learning**

## Discussion

This systematic mapping review provides a broad overview of the available studies on seven motor learning strategies, including their effect on improving functional tasks in neurological and geriatric populations. In total, 87 studies were identified, covering six of the seven included motor learning strategies. The most frequently researched motor learning strategies were dual-task learning (n = 50 studies), mental practice (n = 19), and action observation (n = 12). No studies were found for discovery learning.

### Overview of available studies

Mapping of the publications gave more insight into the development of publications over time and the distribution and quality of RCTs regarding the seven strategies and different target groups. n the early 1990s, new scientific insights were published regarding recovery mechanisms and neuroplasticity of the brain, which fuelled interest in the potential role of motor learning in rehabilitation.^107^ Research on the effects of motor learning strategies to improve functional tasks started around the year 2000, with studies on mental practice and publications increasing substantially from 2010 on. In general, there seems to be quite a big delay between the discovery of a learning strategy’s (potential) working mechanisms and its evaluation in applied research through RCTs. For example, the mirror-neuron system and its role in our ability to learn by imitating others was discovered in the early 1990s,^108,109^ but it took about 20 years before the effects were evaluated within rehabilitation. Comparably, Mellit and Petit^110^ showed through fMRI that during imagery and performance of a motor skill, almost the same brain areas are active, which gave a huge impulse for research on movement imagery in sports. But it wasn’t until 2007 that an increase in studies within (neuro)rehabilitation was seen.

Based on the results of our Delphi study^111^, we expected the number of included studies to be (somewhat) equally balanced across the motor learning strategies. After all, a substantial number of experts had identified these seven motor learning strategies as the ‘most used and well-known’ strategies within their field. This was, however, not the case. By far, dual-task learning has been examined the most. There are several reasons which may explain why. First, dual-tasking is highly prevalent in daily life activities and needs to be practised as such in every context (specificity).^112^

Additionally, some of the first clinical studies conducted showed very promising results (e.g.^23,24,68^) which might have led to an increase in similar, repetitive research paradigms: in 38 of the 50 included studies, walking or balance was trained using a secondary cognitive task. Interestingly, no studies on the use of discovery learning were included (one was excluded during the screening process). And only two studies on analogy learning fulfilled our inclusion criteria.^26,27^ This may be explained by the fact that analogy learning is a relatively new concept, which was first translated to rehabilitation in 2014 for persons after stroke^113^ and with Parkinson’s disease.^26,27,114^ Trial-and-error learning was not researched as the experimental intervention in the included studies but was only used as a control condition for errorless learning in four studies. This is contrary to other fields of research (e.g. children, sports), in which trial-and-error learning and discovery learning have been assessed more extensively.^115^

Although the target population was defined broadly in our search and inclusion criteria, motor learning strategies were studied in only five populations. Older adults (without other specific motor or cognitive problems) were the most researched, followed by persons with Parkinson’s disease and after stroke. No studies were found on other neurological diseases such as multiple sclerosis, spinal cord injury, or traumatic brain injury. Within the included population, certain motor learning strategies appeared to be preferred over others. Errorless learning, for example, was almost exclusively researched in persons with dementia, dual-task learning was mostly studied in older adults, and mental practice was often used in studies to improve arm-hand ability in people after stroke. Based on clinical expertise these strategies may be seen as more suitable for these specific populations, but evidence for their effects is still unclear.

A critical finding of this review is the limited methodological rigour observed across the 87 included studies. Specifically, only five studies (6%) had a low risk of bias and justified *and* achieved their desired sample size. Most of the studies scored ‘some concerns’ (54 studies; 62%). The lack of a strong methodological foundation in many studies makes it difficult to reliably identify the true intervention effects.^116^ We also noted that the interpretation of individual study findings was frequently inaccurate (e.g. solely reported within-group differences and/or interaction effects without reporting between-group differences). There seems to be a more general problem of regulating the risk of bias and ensuring the accuracy of the reporting within motor learning research.^117,118^

### Effects of the seven motor learning strategies

Only four (5% of total) studies were deemed reliable enough to interpret effects reported, based on their RoB2 (category green; low RoB), sample size justification (category green), and reporting of between-group differences: Geroin et al.^77^ found between-group differences for dual-task gait and functional training in persons with Parkinson’s, Trombetti et al.^87^ and Javadpour et al.^81^ found between-group differences for dual-task balance training in older adults, and Jie et al.^26^ did not find significant between-group differences in gait for analogy learning in persons after stroke. Within the remaining moderately reliable studies, there seems to be a clear positive trend favouring observational learning across various populations (8/11 studies), when applied in addition to care as usual. Within mental practice (8/19 studies) and dual-task learning (27/43 studies), between-group effects favouring the intervention group were found in about half and the majority of the included studies, respectively. For dual-task learning, some trends may be observed within populations, as between-group effects in favour of the dual-task interventions were significant for the majority of the studies in persons with Parkinson’s (5/6 studies), dementia (4/5 studies), and older adults with balance impairments (3/3 studies). Remarkably, two studies in dual-task learning in persons with dementia^70^ and older adults with balance impairments,^98^ found between-group differences in favour of the single-task (control) intervention.

### Study limitations

Our search strategy was carefully prepared by experts in the field of literature review and motor learning. Still, we might have missed studies due to limitations in the search strategy (e.g. specific included search terms) and the categorisation of studies within databases. Publication bias might influence our findings; given the diversity of included studies, a funnel plot was not feasible.

As with any review article, our conclusions are subject to some common points of criticism concerning the standardised assessment of the studies’ quality (Minozzi et al.^119^). Despite careful preparation, we still experienced that the use of the RoB2 tool left room for interpretation and needed additional effort to increase the reliability of the assessment. However, missing information in the texts was not retrieved by contacting the authors. Information not reported is not necessarily information not retrieved, and therefore, the criteria list assesses the study’s report, not necessarily the quality of the study.

In line with earlier reviews, we decided to include an additional criterion (i.e. sample size justification) in our assessment of the included studies. An absence of sample size justification is not inherently problematic but increases uncertainties when evaluating effects.^116^ Therefore, we excluded studies with a high risk of bias and without sample size justification from the descriptive analyses.

### Future research

There are several considerations for researchers when conducting applied clinical motor learning research. Researchers should consider evaluating potential rehabilitation strategies not only *within* but also *across* populations.^120^ Despite the anatomic and pathophysiological differences of target populations, these groups share many similarities, ranging from comparable cellular and neuro-physiological responses and recovery mechanisms to the effects of training in motor learning.

We also think that we should reconsider whether RCTs are the best fit for assessing complex training interventions. When conducting RCTs, researchers need to choose between internal validity (e.g. controlled context in laboratory settings) vs external validity reflecting daily practice (e.g. more ‘uncontrolled’ context with potential biases).^121,122^ This may (partly) explain the overall moderate to high risk of bias and perhaps the absence of effects on some occasions. Hence, researchers should consider different research designs, e.g. cohort studies or multiple baseline designs, which might be more suitable for pragmatic trials with complex interventions. To facilitate interpretations of study results by therapists, researchers should also consider using clinically relevant differences, e.g. referring to minimally clinically important differences (MCIDs).^123^ Likewise, to further increase the transferability to clinical practice, careful attention should be given to description of the interventions. The TIDieR (Template for Intervention Description and Replication) can be used as a checklist and guide to ensure interventions are reported with sufficient detail.^124^

### Conclusions

The results of this study provide an overview of the current state of evidence regarding seven motor learning interventions in older neurologic and geriatric rehabilitation. The findings clearly show a skewed distribution of studies across motor learning interventions that have been researched within five target populations. The methodological shortcomings, e.g. high risk of bias and lack of appropriate sample size justifications, make it difficult to draw firm conclusions regarding the effectiveness of motor learning strategies. Hence, this review cannot provide a strong basis for therapists to rely on in their decision-making. Based on observed trends, therapists may consider (to continue) using dual-task learning, observational learning and movement imagery. While waiting for future research, therapists may also consider the other motor learning strategies based on their own experiences and patients’ preferences; the description of the interventions of the included studies could be an example of how to apply different strategies in daily practice within the different neurological and geriatric target populations.

## Data Availability

All data produced in the present study are available upon reasonable request to the authors

## Acknowledgements

We thank Rachel Slangen for her time and expertise in the risk of bias assessment. Furthermore, we also want to show our gratitude to Ciaran Apolo and Jorg van Beek, who helped us extract the data from all the papers during the screening process.

Statement: During the preparation of this work, the authors used the assistance of an AI language model provided by OpenAI, known as ChatGPT, to refine writing. After using this tool, the author(s) thoroughly reviewed and edited the content as needed and take full responsibility for the accuracy and integrity of the public.

This study was funded by Regieorgaan SIA (Dutch Organization for Scientific Research Applied Research Fund) under grant number RAAK.PUB09.001. The funding source had no involvement in the execution of the study or the analysis of the data. The manuscript was registered at international platform of registered systematic review and meta-analysis protocols (INPLASY202430056).

The authors declare no conflict of interest.

