## Appendix 1 for "Overview of effects of motor learning strategies in neurological and geriatric populations: a systematic mapping review"

**Appendix 1:** Search functions used in PubMed, Embase and CINAHL

**PubMed**

|  | **MESH** |  | **Free search terms** |
| --- | --- | --- | --- |
| 1 | ‘Aged’[Mesh] OR ‘Nervous System Diseases’[Mesh] | **OR** | ageing [tiab] OR aging [tiab] OR aged [tiab] OR elderly [tiab] OR old [tiab] OR older [tiab] OR senior* [tiab] OR elder [tiab] OR geriatric [tiab] OR neurological disorder* [tiab] OR brain injur* [tiab] OR left hemisphere [tiab] OR right hemisphere [tiab] OR brain damage [tiab] OR brain lesion [tiab] OR stroke [tiab] OR cva [tiab] OR cerebrovascular accident [tiab] OR brain vascular accident [tiab] OR Parkinson* [tiab] OR dementia [tiab] OR Alzheimer [tiab] OR vascular dementia [tiab] OR Multiple Sclerosis [tiab] OR MS [tiab] |
| **AND** |  |  |  |
| 2 | NA | **OR** | implicit learning [tiab] OR implicit performance [tiab] OR explicit learning [tiab] OR errorless [tiab] OR analogy [tiab] OR analogies [tiab] OR observational learning [tiab] OR action observation [tiab] OR errorful [tiab] OR trial and error [tiab] OR dual task [tiab] OR dualtasking [tiab] OR dual tasking [tiab] OR discovery learning [tiab] OR movement imagery [tiab] OR mental practice [tiab] |
| **AND** |  |  |  |
| 3 | ‘Psychomotor Performance’[Mesh] OR ‘Activities of Daily Living’[Mesh] OR ‘Physical Functional Performance’[Mesh] OR ‘Movement’[Mesh] OR ‘Postural Balance’[Mesh] | **OR** | psychomotor* [tiab] OR motor* [tiab] OR task perform* [tiab] OR ADL [tiab] OR Activities of Daily Living [tiab] OR daily life [tiab] OR walk* [tiab] OR gait [tiab] OR ambulation [tiab] OR mobility [tiab] OR postural control [tiab] OR balance control [tiab] OR locomot* [tiab] OR stand [tiab] OR standing [tiab] OR postural balance [tiab] |
| **AND** |  |  |  |
| 4 | (‘Randomized Controlled Trial’ [Publication Type]) OR ‘Clinical Trial’ [Publication Type] |  | RCT [tiab] OR randomized controlled trial [tiab] OR randomised controlled trial [tiab] OR clinical trial [tiab] |

**CINAHL**

|  | **CINAHL headings** |  | **Free search terms** |
| --- | --- | --- | --- |
| 1 | (MH ‘Aged’) OR (MH ‘Nervous System Dise’ | **OR** | ageing OR aging OR aged OR elderly OR old OR older OR senior* OR elder OR geriatric OR neurological disorder* OR brain injur* OR left hemisphere OR right hemisphere OR brain damage OR brain lesion OR stroke OR cva OR cerebrovascular accident OR brain vascular accident OR Parkinson* OR dementia OR Alzheimer OR vascular dementia OR Multiple Sclerosis OR MS |
| **AND** |  |  |  |
| 2 | NA | **OR** | implicit learning OR implicit performance OR explicit learning OR errorless OR analogy OR analogies OR observational learning OR action observation OR errorful OR trial and error OR dual task OR dualtasking OR dual tasking OR discovery learning OR movement imagery OR mental practice |
| **AND** |  |  |  |
| 3 | (MH ‘Psychomotor Performance’) OR (MH ‘Physical Performance’) OR (MH ‘Activities of Daily Living’) OR (MH ‘Activities of Daily Living (Saba CCC)’) OR (MH ‘Movement’) OR (MH ‘Balance, Postural’) | **OR** | psychomotor* OR motor* OR task perform* OR ADL OR Activities of Daily Living OR daily life OR walk* OR gait OR ambulation OR mobility OR postural control OR balance control OR locomot* OR stand OR standing OR postural balance |
| **AND** |  |  |  |
| 4 | (MH ‘Randomized Controlled Trials’) OR (MH ‘Clinical Trials’) |  | RCT OR randomized controlled trial OR randomised controlled trial OR clinical trial |

**Embase**

|  | **Emtree** |  | **Free search terms** |
| --- | --- | --- | --- |
| 1 | 'aged'/exp OR 'geriatric disorder'/exp OR 'central nervous system disease'/exp OR 'dementia'/exp | **OR** | ageing OR aging OR aged OR elderly OR old OR older OR senior* OR elder OR geriatric OR neurological disorder* OR brain injur* OR left hemisphere OR right hemisphere OR brain damage OR brain lesion OR stroke OR cva OR cerebrovascular accident OR brain vascular accident OR Parkinson* OR dementia OR Alzheimer OR vascular dementia OR Multiple Sclerosis OR MS |
| **AND** |  |  |  |
| 2 | NA | **OR** | implicit learning OR implicit performance OR explicit learning OR errorless OR analogy OR analogies OR observational learning OR action observation OR errorful OR trial and error OR dual task OR dualtasking OR dual tasking OR discovery learning OR movement imagery OR mental practice |
| **AND** |  |  |  |
| 3 | 'psychomotor performance'/exp OR 'physical performance'/exp OR 'daily life activity'/exp OR 'movement (physiology)'/exp OR 'body equilibrium'/exp | **OR** | psychomotor* OR motor* OR task perform* OR ADL OR Activities of Daily Living OR daily life OR walk* OR gait OR ambulation OR mobility OR postural control OR balance control OR locomot* OR stand OR standing OR postural balance |
| **AND** |  |  |  |
| 4 | 'randomized controlled trial'/exp OR 'clinical trial'/exp |  | RCT OR ‘randomized controlled trial’ OR ‘randomised controlled trial’ OR ‘clinical trial’ |
